# Cardiomyocyte-derived circulating extracellular vesicles allow a non-invasive liquid biopsy of myocardium in health and disease

**DOI:** 10.1101/2024.09.19.24314009

**Authors:** Michail Spanos, Priyanka Gokulnath, Guoping Li, Elizabeth Hutchins, Bessie Meechoovet, Quanhu Sheng, Emeli Chatterjee, Ritin Sharma, Natacha Carnel-Amar, Claire Lin, Christopher Azzam, Ima Ghaeli, Kaushik V Amancherla, José Fabian Victorino, Krystine Garcia-Mansfield, Ryan Pfeffer, Parul Sahu, Brian R. Lindman, Sammy Elmariah, Eric R. Gamazon, Michael J. Betti, Xavier Bledsoe, Michelle L. Lance, Tarek Absi, Yan Ru Su, Ngoc Do, Marta Garcia Contreras, Dimitrios Varrias, Michail Kladas, Miroslav Radulovic, Dimitris Tsiachris, Anastasios Spanos, Konstantinos Tsioufis, Patrick T. Ellinor, Nathan R. Tucker, James L. Januzzi, Patrick Pirrotte, Tijana Jovanovic- Talisman, Kendall Van Keuren-Jensen, Ravi Shah, Saumya Das

**Affiliations:** Cardiovascular Research Center, Massachusetts General Hospital and Harvard Medical School, Boston, MA, USA; Division of Neurogenomics, The Translational Genomics Research Institute, Phoenix, AZ, USA; Laboratory of Neurogenetics, National Institute on Aging, National Institutes of Health, Bethesda, MD, USA; Department of Biostatistics (Q.S.), Vanderbilt University Medical Center, Nashville, TN; Collaborative Center for Translational Mass Spectrometry, Translational Genomics Research Institute, Phoenix, AZ, USA; Cancer & Cell Biology Division, Translational Genomics Research Institute, Phoenix, AZ; Department of Molecular Medicine, Beckman Research Institute, City of Hope Comprehensive Cancer Center, Duarte, CA, USA; Masonic Medical Research Institute, Utica, NY, USA 13501; Vanderbilt Translational and Clinical Research Center (R.S.), Vanderbilt University Medical Center, Nashville, TN, USA; Division of Cardiology, Massachusetts General Hospital and Harvard Medical School, Boston, MA, USA; Department of Medicine, Cardiovascular Division, University of California-San Francisco, San Francisco, CA, USA; Vanderbilt Genetics Institute, Vanderbilt University Medical Center, Nashville, TN 37203, USA; SUNY Upstate Medical University, Syracuse, New York, USA; Department of Cardiac Surgery, Vanderbilt University Medical Center, Nashville, Tennessee, USA; Spectradyne LLC, Signal Hill, CA, USA; Albert Einstein College of Medicine/ Jacobi Medical Center, The Bronx, NY, USA; Albert Einstein College of Medicine/ North Central Bronx/Jacobi Medical Center, New York City Health and Hospitals, The Bronx, NY, USA; First Department of Cardiology, School of Medicine, National and Kapodistrian University of Athens, Hippokration General Hospital, Athens, Greece; Department of Cardiology, Athens Medical Center, Athens, Greece; Baim Institute for Clinical Research, Boston, Massachusetts, USA

## Abstract

The ability to track disease without tissue biopsy in patients is a major goal in biology and medicine. Here, we identify and characterize cardiomyocyte-derived extracellular vesicles in circulation (EVs; “cardiovesicles”) through comprehensive studies of induced pluripotent stem cell-derived cardiomyocytes, genetic mouse models, and state-of-the-art mass spectrometry and low-input transcriptomics. These studies identified two markers (*POPDC2*, *CHRNE*) enriched on cardiovesicles for biotinylated antibody-based immunocapture. Captured cardiovesicles were enriched in canonical cardiomyocyte transcripts/pathways with distinct profiles based on human disease type (heart failure, myocardial infarction). In paired myocardial tissue-plasma from patients, highly expressed genes in cardiovesicles were largely cardiac-enriched (vs. “bulk” EVs, which were more organ non-specific) with high expression in myocardial tissue by single nuclear RNA-seq, largely in cardiomyocytes. These results demonstrate the first “liquid” biopsy discovery platform to interrogate cardiomyocyte states non-invasively in model systems and in human disease, allowing non-invasive characterization of cardiomyocyte biology for discovery and therapeutic applications.

## INTRODUCTION

Through the last two decades, precision clinical risk assessment and phenotyping in cardiovascular disease (CVD) has benefited from efforts to rapidly and simultaneously profile biomolecules from DNA to physiologic effectors (“omics”) in at-risk populations^1–8^. While “omic” approaches to precision CVD have derived primarily from plasma^6,8^, an increasing literature underscores the importance of tissue phenotypes in risk stratification and discovery, particularly in highly heterogeneous states like aging and heart failure (HF)^9^. Recent efforts to index plasma molecular profiles to tissue rely heavily on tissue atlases^9^—derived from limited populations not specific to CVD—which therefore lack sensitivity for the shifting molecular and clinical landscape during disease progression. Indeed, these atlas-based approaches have found well-established biomarkers that reflect the “end-phenotypes” of HF (troponin, natriuretic peptides^10–12^), limiting the discovery of dynamic, upstream <u>targetable</u> pathways that may unlock precision phenotyping of disease heterogeneity for targeting. Unlike cancer, where tissue diagnostics are required for targeted treatment^13,14^, obtaining myocardial tissue remains appropriately challenged by ethical constraints and risk. Identifying a circulating, <u>tissue-enriched</u> molecular signature that captures a myocardial state at an <u>individual</u> level quantitatively, precisely, and serially will sharpen precision from broad “bulk” plasma molecular profiling to improve risk stratification, disease sub-type identification, and novel target discovery.

Extracellular vesicles (EVs) are membrane-delimited particles secreted from every known cell type that carry molecular cargo (proteins, metabolites, transcripts), reflecting the composition of their source cell^15–17^. Here, we propose a translational framework to identify, characterize, and profile a population of cardiomyocyte-enriched EVs across CVD states, including HF. We selected EV as a potential source of RNA biomarkers representative of their cardiomyocyte source, given known release from the heart during metabolic stress^18^ as well as prior studies suggesting that EV molecular cargo may harbor transcriptional signatures relevant to human HF^19^. The overall scheme of our study is summarized in **Fig. 1**. Our multi-parametric approach used mass spectrometry proteomics to identify membrane proteins on EVs from induced pluripotent stem cell (iPSC)-derived cardiomyocytes, winnowing targets via additional computational approaches, atlases, and based on performance of antibodies against targets. We selected two surface proteins, *POPDC2* and *CHRNE,* specific for cardiomyocyte-derived EVs (termed “cardiovesicles”), developing an immunoaffinity-based pull-down technique (EV immunocapture methodology termed “Exocapture-MSB”) to capture and characterize cardiovesicles using antibodies against these two targets from plasma. Our work subsequently characterized the transcriptional cargo of these cardiovesicles in a genetically engineered mouse model system tagging cardiac-derived EVs for capture as well as human plasma from patients with and without CVD. We finally used paired myocardial tissue-plasma from patients to assess similarities between top expressed transcripts in plasma cardiovesicles and myocardial tissue (single nuclear RNA-seq). This multi-modality translational demonstration of cardiomyocyte-derived EVs offers a new paradigm of non-invasive myocardial phenotyping currently inaccessible to bulk “omic” studies and outlines a discovery paradigm for implementation across multiple organ systems. Process optimization and, ultimately, clinical application of these methodologies may open up new avenues for precision medicine approaches to cardiovascular risk stratification and targeted treatment.

**Figure 1.**
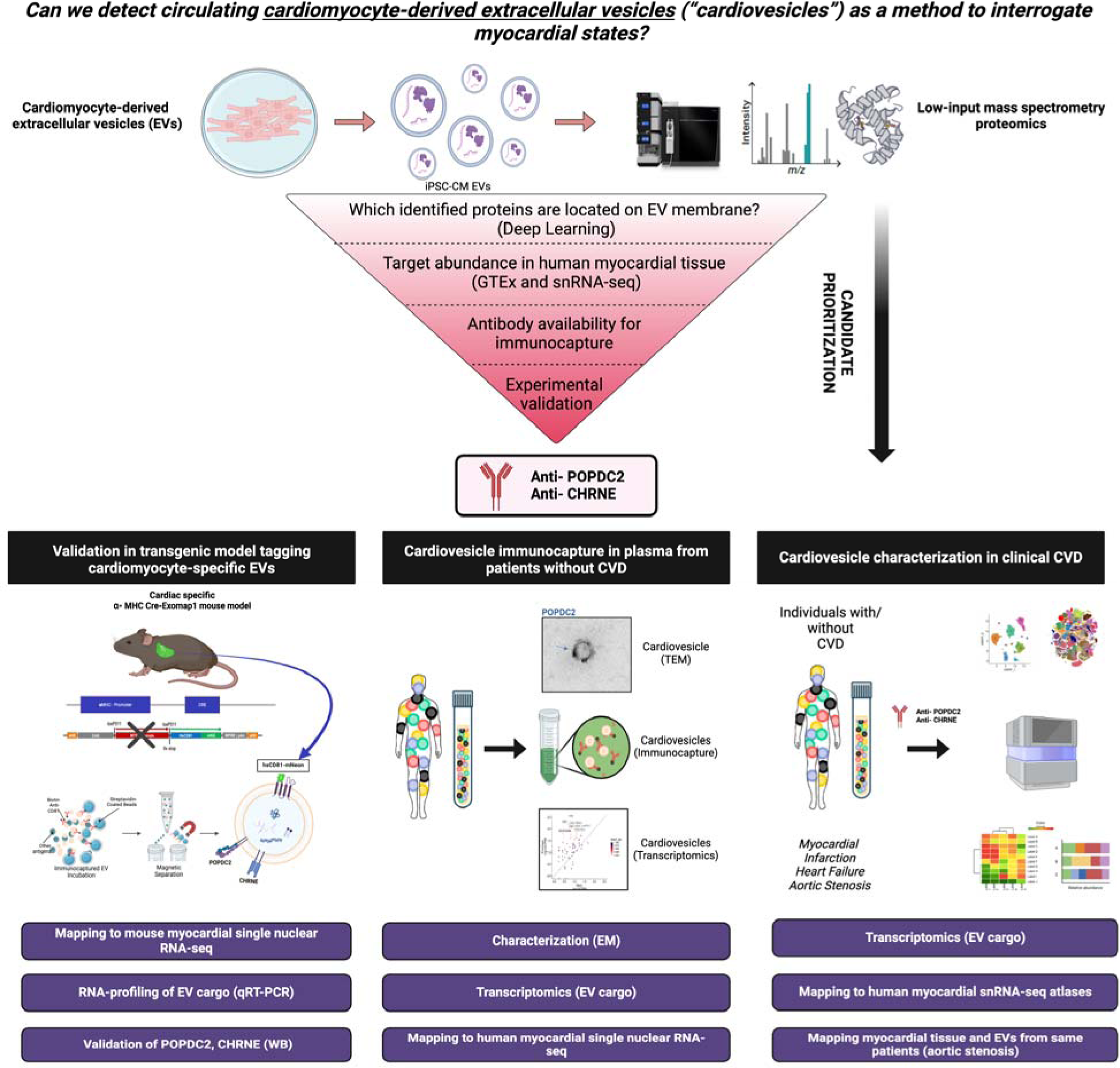
Study design to identify cardiovesicles for liquid biopsy in cardiovascular diseases. Proteomics and bioinformatics-based approach to identify POPDC2 and CHRNE as cardiovesicular membrane proteins with validation in human RNA-seq datasets and in vitro experiments. Confirmation of cardiomyocyte EV-specific expression of POPDC2 and CHRNE proteins using the Exomap1 transgenic mouse model (humanized CD81 expression restricted to cardiomyocytes and its EVs). Characterization of the RNA cargo of cardiovesicles in healthy human plasma. Cardiovesicle RNA cargo characterization in cardiovascular disease cohorts with mapping to tissue single cell RNA sequencing data sets.

## RESULTS

### Identifying membrane protein markers to enable cardiomyocyte-derived EV isolation

**Fig. 2a** outlines our analytic scheme to identify cardiomyocyte-specific surface protein expression on EVs using two complementary approaches. ***In vitro* proteomic approach**: we first used a proteomics-based approach in iPSC-derived cardiomyocytes (iPSC-CMs) (**Fig. S1a, b**) and their released EVs (iPSC-CM-EVs) to identify proteins expressed within cardiomyocytes <u>and</u> on the surface of EVs derived from those cells. We established the following criteria for our protein discovery process: 1) expression in both iPSC-CMs and iPSC-CM-EVs, 2) elevated levels in human heart tissues, and 3) transmembrane expression. Of the 5,953 proteins measured by liquid chromatography-mass spectrometry (5,397 in iPSC-CMs; 4,647 in iPSC-CM-EVs, **Fig. S1c, Table S1**), 4,091 were shared by both (**Fig. S1d**). Among these, 150 were enriched in cardiac tissue according to the Human Protein Atlas^20^ (HPA, **Fig. S1e**). For effective downstream capture of EVs, it is crucial that the identified proteins have an exposed portion and are transmembrane. Consequently, we employed computational subcellular localization algorithms (Deep Transmembrane Helix Prediction - DeepTMHMM^21^) to identify those with transmembrane structures. This analysis refined our initial selection from 150 to 35 cardiac-enriched membrane proteins that met all criteria, including transmembrane expression in both iPSC-CMs and iPSC-CM-EVs with elevated levels in human heart tissues (**Fig. 2b**). By prioritizing those with an increased combined abundance score in both iPSC-CMs and iPSC-CM-EVs and excluding non-specific mitochondrial proteins, we identified the top 10 most abundant candidate proteins (ATP2A2, SLC25A4, SLC25A11, CDH2, NCAM1, RYR2, PLN, SLC8A1, POPDC2, and SLC27A6).

**Figure 2.**
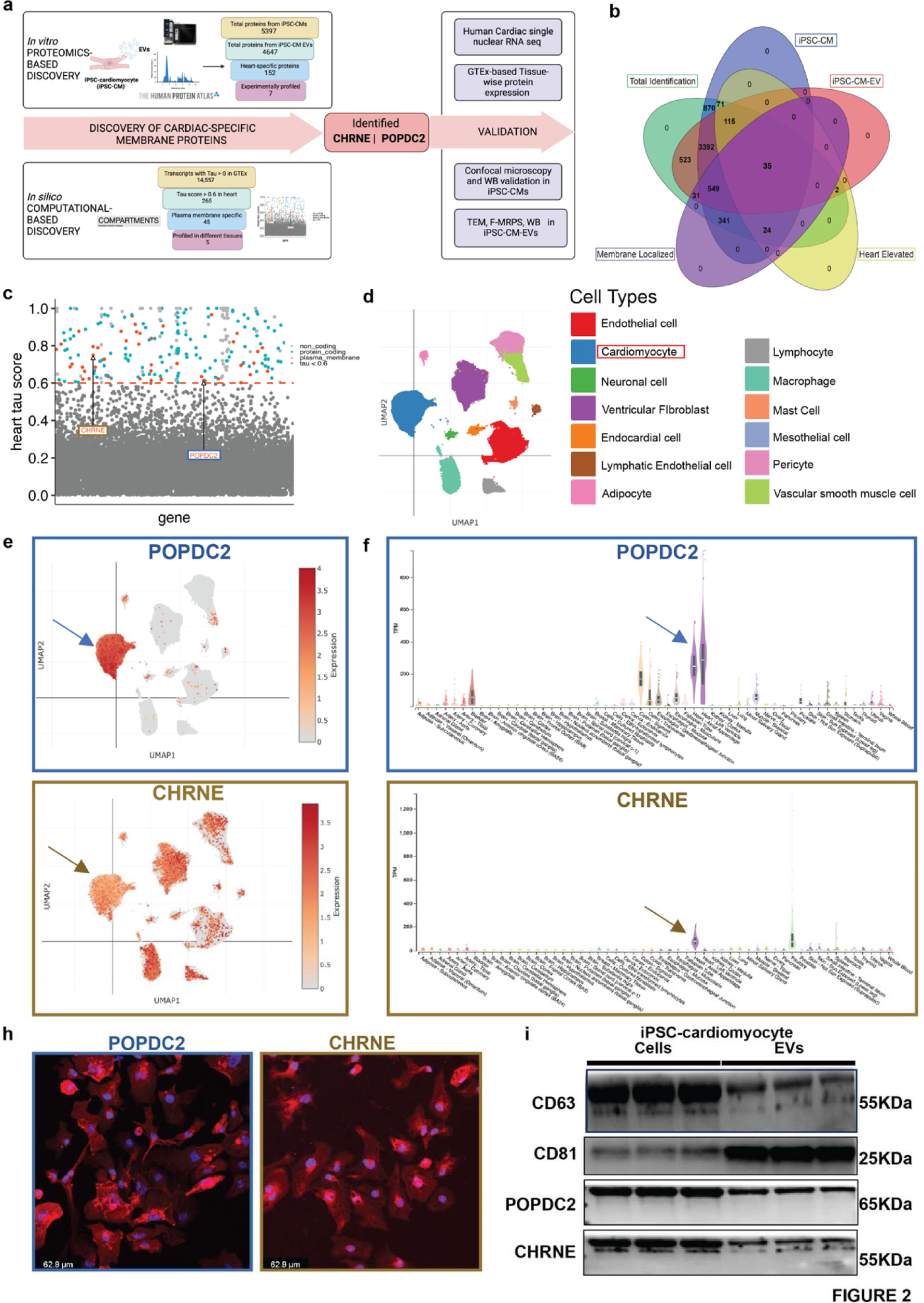
Discovery and experimental validation of POPDC2 and CHRNE as cardiomyocyte-EV (Extracellular Vesicle) membrane protein candidates. **a.** Analytic scheme for identification and validation of cardiomyocyte EV membrane markers POPDC2 and CHRNE using step-wise proteomic-based and bioinformatic-based discovery followed by experimental validation in iPSC-CM cells and EVs. **b.** Venn diagram showing 35 targets that were identified using LC-MS/MS (Liquid Chromatography with tandem mass spectrometry) present in iPSC-CM (induced Pluripotent Stem Cell-derived Cardiomyocytes), iPSC-CM-EVs and prioritized by heart enrichment (from Human Protein Atlas data) and membrane-localization (Deep Transmembrane Helix Prediction). **c.** Scatter plot showing candidates from computational analysis obtained by mining GTEx for proteins with cardiac-specificity (tau) ≥ 0.6 (dots above the red dashed line) with red indicating protein-coding candidates present in the plasma membrane (predicted using Compartments). POPDC2 and CHRNE are indicated on the plot. **d.** UMAP (Uniform Manifold Approximation and Projection) legend of cardiac single nuclear RNA-seq dataset from Broad single cell Portal. **e.** POPDC2 and CHRNE UMAPs demonstrating their expression in cardiomyocytes. **f.** Tissue-wise transcriptomic expression of POPDC2 and CHRNE using GTEx showing elevated expression in heart tissue. **h.** Representative confocal images showing the expression of POPDC2 and CHRNE proteins in iPSC-CMs. **i**. POPDC2 and CHRNE protein expression shown in iPSC-CMs and iPSC-CM-EVs by western blotting (n=3 independent experiments).

### *In silico* computational approach

Given the known limitation of iPSC-CMs in modeling human adult CMs^22^, we conducted an *in silico* tissue specificity analysis to prioritize targets for antibody selection in parallel (**Fig. 2c**). Within GTEx, we calculated a tau specificity score (see **Methods**) for each RNA (long non-coding and protein-coding RNAs) to determine tissue-specificity of expression. By including lncRNA in this analysis, in addition to using these data to prioritize candidates, we are also able to evaluate the RNA cargo in the resulting EV populations for tissue specificity using both protein coding genes and lncRNA. Of the 14,557 genes with an assigned tau value > 0, there are 265 genes with a tau score in the heart >= 0.6 (**Fig. 2c**; dots above the horizontal dashed line), indicating significant enrichment in heart tissue at a transcriptional level. Of these 265, 173 were protein-coding (**Fig. 2c**; blue and red dots). Similarly to the *in vitro* proteomic approach, to ensure that we prioritized membrane proteins (to enable subsequent immunocapture), we characterized subcellular localization of gene products via the Compartments database^23^. This computational approach identified 45 cardiac-enriched plasma membrane proteins (heart tau score ≥ 0.6; **Fig. 2c**, red dots), including CHRNE and POPDC2.

### Target prioritization

The final 55 candidate proteins identified by the above approaches (10 from the in vitro proteomic and 45 from the in silico computational approach) underwent testing for immunocapture potential by (1) experimental screening for expression in iPSC-CMs and iPSC-CM-EVs, (2) assessing cardiac specificity by testing across various human tissues, (3) evaluating availability and efficacy of commercial antibodies and suitability for biotinylation. Representative examples of this experimental validation step to prioritize the targets are shown in **Fig. S1f, g**. POPDC2 and CHRNE consistently passed these filters, demonstrating tissue enrichment in GTEx at a transcriptional and proteomic (for POPDC2) level (**Fig. S1h**). In addition, using published myocardial single nuclear RNA-seq data^24^, we demonstrated predominant expression of POPDC2 and CHRNE in myocardial tissue at a cellular level (**Fig. 2d-h**). Cellular localization of POPDC2 and CHRNE in iPSC-CMs was verified via confocal microscopy (**Fig. 2h**), and expression in iPSC-CM-EVs was confirmed by Western blot (WB) (**Fig. 2i**), Transmission Electron Microscopy (TEM), and flow cytometry (**Fig. S1i, j**). Finally, we were able to validate commercially available antibodies against these two antigens both in immunoblotting (WB) (**Fig. 2i**) and subsequent immunocapture experiments. Given these multiple lines of corroborating humanized cellular, tissue, informatic, and morphologic evidence, we selected POPDC2 and CHRNE to carry forward as targets for antibody-mediated pull-down of cardiomyocyte-enriched EVs.

### POPDC2-/CHRNE-bearing EVs from plasma have a cardiomyocyte origin *in vivo*

Before moving to human translational studies, we next sought to confirm that POPDC2- and CHRNE-studded EVs in plasma were of cardiomyocyte origin *in vivo*. We utilized a genetically engineered mouse model (ExoMAP1^25^) to track and characterize cardiomyocyte-specific EVs (**Fig. 3a**). This model incorporates a *cre-lox* genetic “switch” system with deletion of TdTomato and expression of a humanized version of CD81 (hsCD81) tagged with monomeric Neon green (mNG) in *Cre*-recombinase-expressing cells. Crossing ExoMAP1 mice with α-myosin heavy chain (MHC)-*Cre* transgenic mice results in progeny with cardiomyocyte-restricted Cre expression, effectively restricting hsCD81/mNG expression to cardiomyocytes and cardiomyocyte-derived EVs (**Fig. 3b**). This genetic construct allowed us to trace cardiomyocyte-derived EVs with precision.

**Figure 3.**
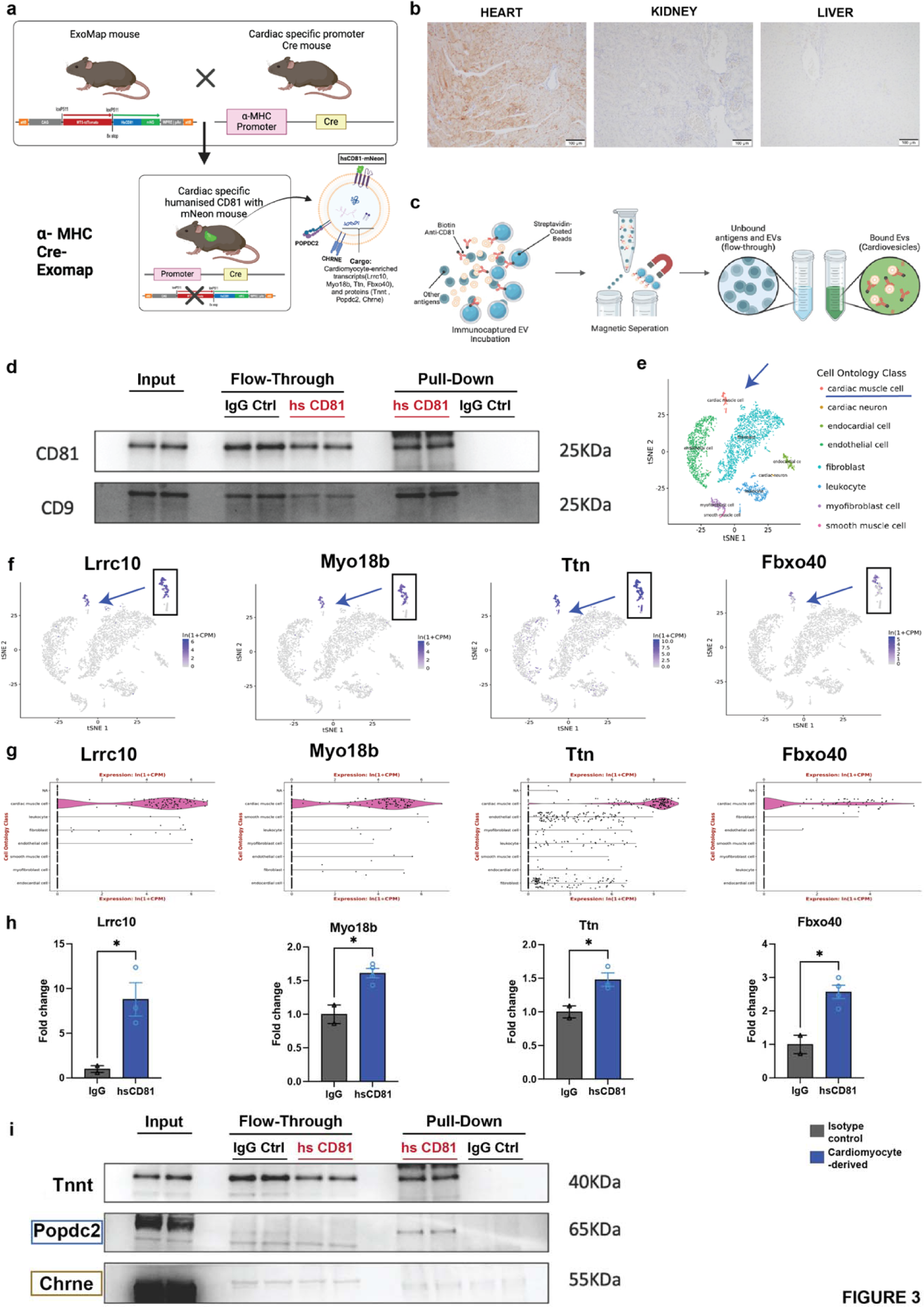
Cardiomyocyte-EV Specific Expression of POPDC2 and CHRNE Proteins in Cardiac-Specific Cre-Driven EXOMAP1 Mouse Model. **a.** Schematic detailing the generation of cardiac-specific EXOMAP1 transgenic mice. Exomap1 mice, which express HsCD81mNG (humanized CD81 fused with mNeonGreen) in a Cre recombinase-dependent manner, were crossed with alpha myosin heavy chain (αMHC)-Cre mice, ensuring HsCD81mNG expression on cardiomyocyte membranes and their secreted EVs. **b.** Cardiac-specific hsCD81 expression as observed in immunohistochemistry staining of heart (positive expression), and absent in kidney and liver**. c.** Overview of the ExoCapture-MSB method for EV immunocapture using biotin-streptavidin affinity and Streptavidin Magnetic Beads targeting cardiac-tissue specific EVs from αMHC-Cre Exomap1 mice. **d.** Confirmation of canonical EV markers CD81 and CD9 enrichment in cardiac-derived EVs via Western blot assay, with controls (PD: Pulled-Down, FT: Flow-Through). **e.** UMAP legend of cardiac single nuclear RNA-seq data from Tabula muris. **f.** and **g.** UMAPs and Violin plots demonstrating enrichment of cardiomyocyte-specific transcripts Lrrc10, Myo18b, Ttn, and Fbxo40 from Tabula muris. **h.** Bar plots showing significant differences in cardiomyocyte-specific transcripts between humanized CD81-positive EV pulldown and isotype control, with respective p-values using Mann Whitney t-test (Lrrc10, p = 0.0482; Myo18b, p=0.0105; Ttn p=0.0459; FBxo40-p=0.0103) n= 4 mice. **i.** Confirmation of POPDC2, CHRNE, and TNNI3 enrichment in cardiac-derived EVs via Western blot assay, validating the selective presence of these key cardiomyocyte markers, affirming their cardiac origin. N= 4 mice.

To capture cardiomyocyte-derived hsCD81^+^ EVs, we developed and employed an immune-affinity-based pull-down method specifically optimized for EV immunocapture, the ExoCapture-MSB technique, which uses streptavidin-coated paramagnetic microspheres combined with high-affinity biotin-streptavidin binding (**Fig. 3c**; see **Methods** for details of the ExoCapture-MSB methodology). We confirmed the successful immunocapture of these EVs relative to isotype control using Western blot assays (**Fig. 3d**). After successful isolation of hsCD81^+^ EVs in our genetic model, we next assessed the cardiac-specificity of their cargo via proteomic (WB) and transcriptomic (qPCR) studies (**Fig. 3f-i**).

Murine cardiomyocyte-enriched transcripts (expected to be more abundant in EVs sourced from cardiomyocytes) were prioritized by their heart-tau scores, expression in Mouse ENCODE transcription data (prioritized by highest RPKM expression in heart with respect to other tissues, **Fig. S2a, b**), and enriched expression in cardiomyocytes from the mouse heart UMAPs obtained from the Tabula muris^26^ database (**Fig. 3e-g**). Quantitative RT-PCR analysis confirmed that the immunocaptured hsCD81^+^ EVs contained cardiac-specific transcripts, including *Lrrc10* (Tau-1), *Myo18b* (Tau-0.763), *Ttn* (Tau-0.267), and *Fbxo40* (Tau-0.952), compared to control IgG (non-specific) immunocapture (**Fig. 3h**). Additionally, the microRNA miR-30d-5p, known for its cardiomyocyte origin and role in cardiac disease^27^, exhibited significant enrichment in immunocaptured hsCD81^+^ EVs relative to IgG control (**Fig. S2c**), consistent with previous findings in iPSC-CMs (**Fig. S3)**. Western blotting confirmed the presence of cardiac troponin, a cardiomyocyte-specific sarcomeric protein, in the hsCD81^+^ EVs (**Fig. 3i**), further corroborating their cardiomyocyte origin.

Finally, after validating the fidelity of our genetic mouse model to capture cardiomyocyte-derived EVs, we demonstrated the membrane enrichment of POPDC2 and CHRNE on cardiomyocyte-derived hsCD81^+^ EVs (**Fig. 3i**). As an appropriate negative control, we isolated hsCD81^+^ EVs from the plasma of mice derived from crossing *vav-cre* transgenic mice (cre expression in hematopoietic cells^28^) (**Fig. S2d**) and ExoMap1 mice. As expected, these EVs were enriched for immune markers (CD45), but not POPDC2 or CHRNE. Together, our in vivo data validated POPDC2 and CHRNE as suitable targets for proceeding with the immunocapture of cardiomyocyte-derived EVs in human studies.

### Cardiomyocyte-derived EVs from human plasma demonstrate cardiac-specific gene expression profiles

We next isolated and characterized POPDC2- and CHRNE-studded EVs (now termed “cardiovesicles”) from pooled plasma of individuals without active cardiovascular disease (**Fig. 4a-b, Fig. S3a-d**). Circulating plasma EVs were isolated by size exclusion chromatography, and were confirmed to contain canonical EV surface markers (CD81, CD9). We demonstrated the presence of POPDC2 and CHRNE on the circulating plasma EVs via multiple gold-standard methods, including (1) TEM with gold-particle-conjugated antibodies, (2) fluorescence microfluidic resistive pulse sensing (MRPS, **Fig. 4a-b**), and (3) high-throughput flow cytometry with microscopic imaging (confirming localization of POPDC2 and CHRNE in human plasma-derived EVs; **Fig. S4d**). Notably, we observed cardiovesicles that were marked by: i) POPDC2 alone; ii) CHRNE alone; iii) both CHRNE and POPDC2. MRPS suggested a higher abundance of POPDC2-marked (POPDC2^+^) cardiovesicles in the plasma (**Fig. 4a**, blue arrow for POPDC2 and gold arrows for CHRNE). Using the ExoCapture-MSB method in human plasma to isolate cardiovesicles (**Fig. 4c**), we tailored and optimized isolation for POPDC2 and CHRNE to maximize immunocapture efficiency (optimization and bead selection). POPDC2- or CHRNE-immunocapture from human plasma yielded cardiovesicles enriched for cardiac-specific troponin (**Fig. 4d-e**), and transcriptome-wide assessment of gene expression in both CHRNE^+^ and POPDC2^+^ cardiovesicles by RNAseq demonstrated largely cardiac-enriched genes relative to bulk plasma EVs (as measured by tissue-enrichment heart tau ≥ 0.9 score from GTEx; **Fig. 4f-g**). Consistent with our double transgenic mouse model, specific cardiac transcripts (*BMP10*, *TNNI3*, *LRRC10*, *FBXO40*) were enriched within cardiovesicles relative to bulk plasma EVs (**Fig. 4h, Fig. S4e-g**). We observed that both POPDC2^+^ and CHRNE^+^ cardiovesicles had consistently higher enrichment of these transcripts. These results were confirmed via a cardiac single nuclear RNA -seq dataset^24^, where transcripts highly expressed in cardiovesicles demonstrated cardiomyocyte selectivity (**Fig. 4i**). These findings strongly suggested the translatability of POPDC2 and CHRNE immunocapture to generate a population of cardiomyocyte-enriched EVs for study.

**Figure 4.**
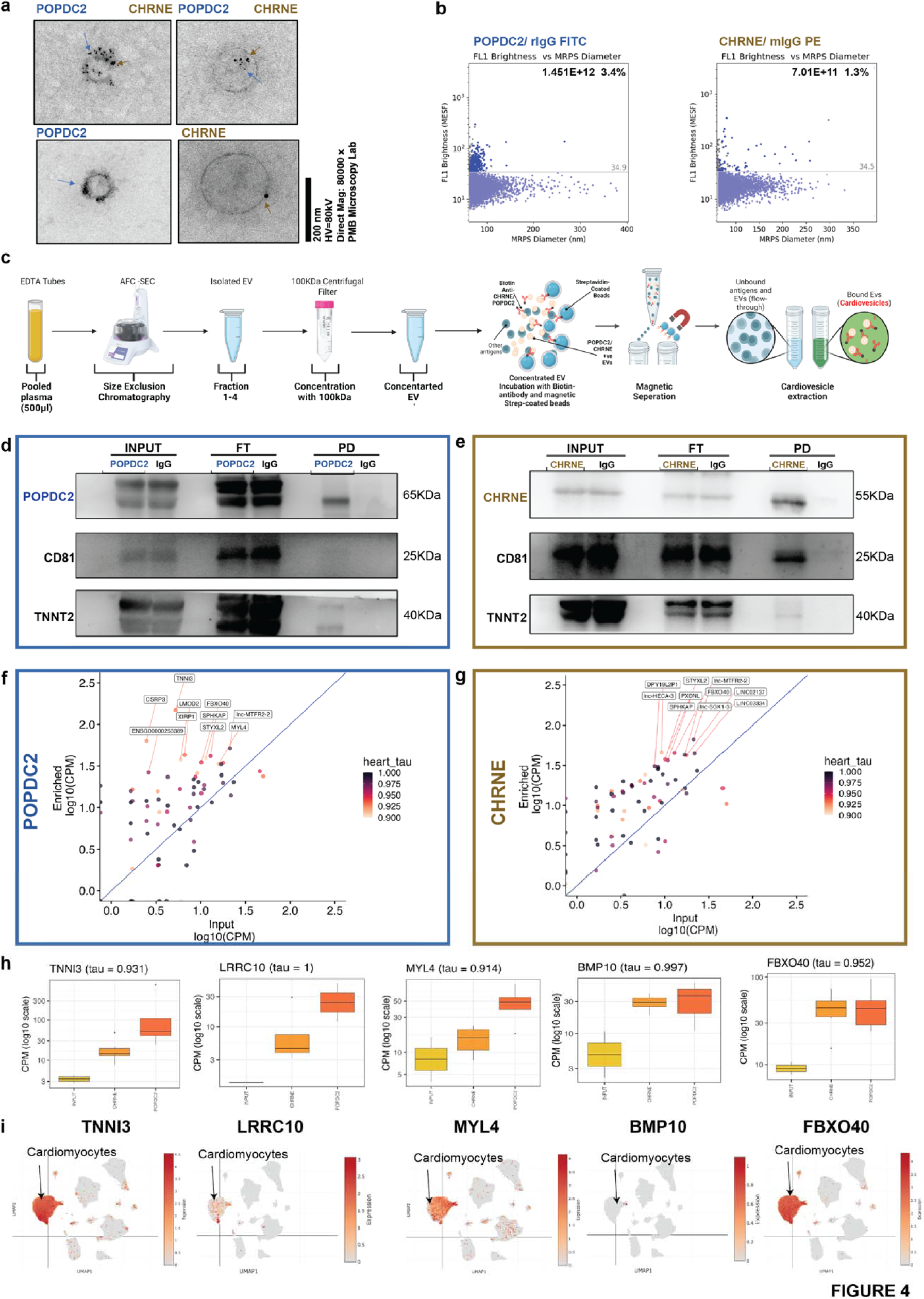
POPDC2+ and CHRNE+ Cardiovesicles from Human Plasma Demonstrate Cardiac-Specific Gene Expression Profiles. **a.** Transmission Electron Microscopy images of POPDC2+ (blue arrows) and CHRNE+ (gold arrows) EVs in healthy human plasma, displaying both single positive (POPDC2+ CHRNE-; POPDC2-CHRNE+) and double positive (POPDC2+ CHRNE+) EVs, with gold particle sizing for POPDC2 (5 mm) and CHRNE (12 mm). **b.** Cardiovesicles exhibiting POPDC2+ and CHRNE+ signatures are visualized and quantified using Fluorescent-Microfluidic Resistive Pulse Sensing (F-MRPS) in plasma EVs (n=3 replicates)**. c.** Detailed overview of the ExoCapture-MSB Method for immunocapture of cardiovesicles from 500 µL of Human Plasma, employing Pierce™ Streptavidin Magnetic Beads with biotin-streptavidin affinity. Western Blot analysis of POPDC2 (**d)** and CHRNE (**e**) immunocapture, highlighting enrichment for cardiomyocyte-specific protein Troponin. Transcriptomic enrichment of cardiac-specific transcripts, prioritized by tau score ≥ 0.9, in vesicles immunocaptured with Biotinylated POPDC2 (**f**) and CHRNE (**g**) antibody, respectively. with the expression of the bulk CD81 immunocaptured vesicles (Input) on the x-axis and the expression of the immunocaptured Cardiovesicles on the y-axis (Enriched). **h.** Box plots showing the expression of heart enriched genes (TNNI3, LRRC10, MYL4, BMP10, FBXO40) in vesicles immunocaptured with CD81, POPDC2, and CHRNE antibodies, confirming specific capture and analysis of Cardiovesicles. **i.** UMAP visualization from single nuclear RNA sequencing depicting plots (TNNI3, LRRC10, MYL4, BMP10, FBXO40) as highly cardiac-enriched transcripts within Cardiovesicles, affirming their cardiac origin. P<0.05 is utilized as the threshold of statistical significance. N= 3 replicates.

### Plasma cardiovesicles capture myocardial disease state-specific expression profiles in patients

We next assessed whether cardiovesicles could represent a source of pathologic molecular information across cardiovascular disease states in patients. We systematically isolated POPDC2- and CHRNE-bearing cardiovesicles and characterized their RNA cargo from 45 patients (15 prospectively recruited with heart failure with reduced ejection fraction; HF, 15 with myocardial infarction; MI; **Fig. 5a**; clinical characteristics in **Tables S2-3**) using EV-RNA sequencing. Cardiovesicle RNA-seq detected 72,503 transcripts, of which 19,494 were protein-coding shared across all groups, which likely reflect RNA transcripts present in EVs independent of disease conditions (such as housekeeping genes). Of the 19,402 transcripts detected in the CHRNE group and 19,442 in the POPDC2 group across all conditions, 99% (19,362) were shared between CHRNE and POPDC2. To confirm the presence of highly cardiac-specific genes in the cardiovesicles, we applied a filter for cardio-specificity score (Tau ≥0.6 from GTEx data), identifying 173 highly specific genes. Of these, 169 were again detected across both cardiovesicle populations (**Fig. 5b**), with the similarity of their cargoes supporting the cardiomyocyte origin of both types of cardiovesicles. When we analyzed the most abundant transcripts within POPDC2^+^ cardiovesicles (top 500 most expressed transcripts prioritized by their average normalized count across all 45 samples), 234 transcripts were common between all three groups, while there were 134 and 119 unique genes in the cardiovesicles isolated from the patients with HF and MI, respectively when compared to the control group, suggesting unique transcriptional patterns in these disease states (**Fig. 5c**). Similarly, 228 transcripts were common in all CHRNE^+^ cardiovesicle groups, with 137 unique for MI and 155 specifically expressed in the HF group (**Fig. 5d**; All results pertaining to CHRNE can be found in the **Fig. S5**).

**Figure 5.**
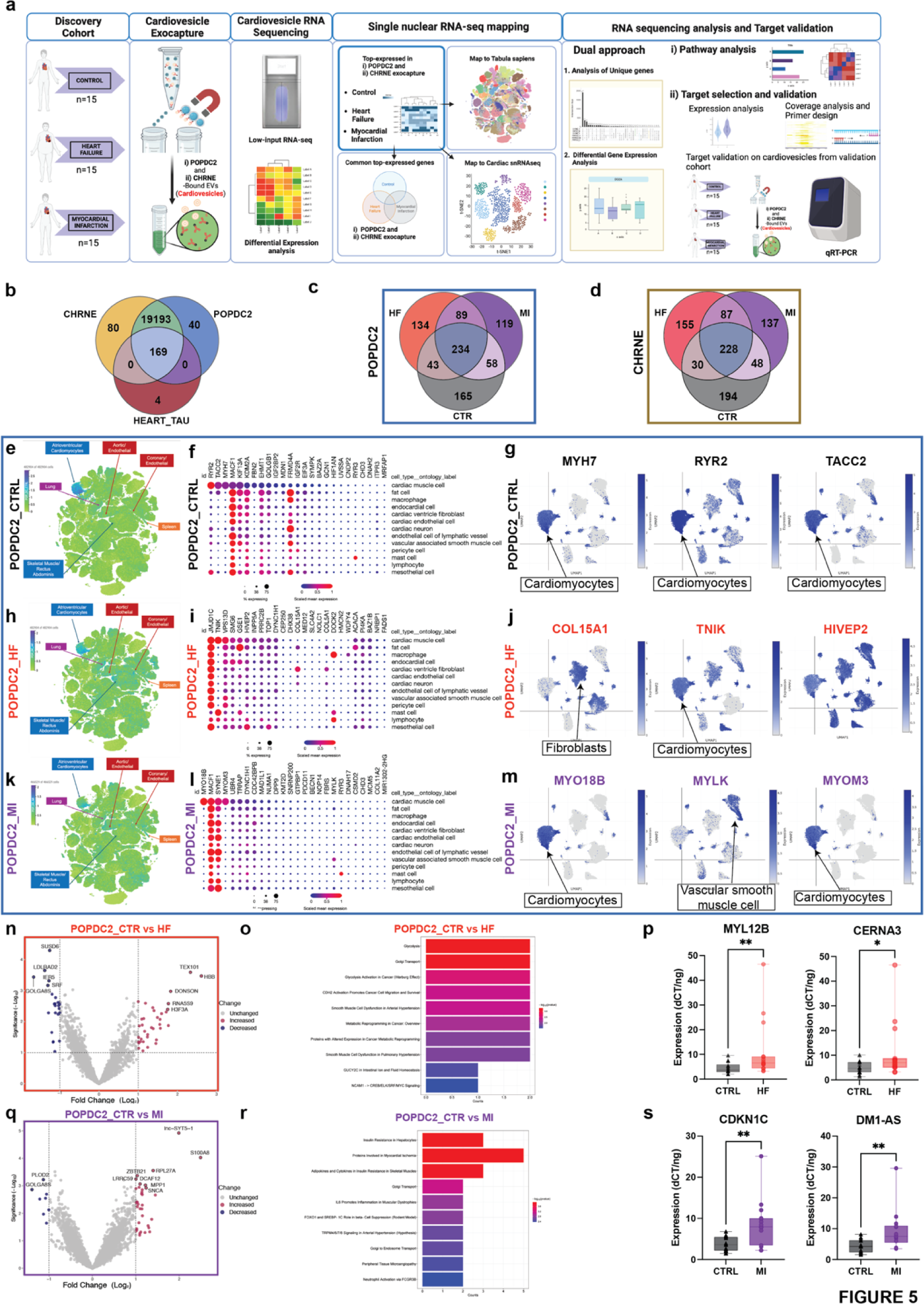
Cardiovesicle transcripts capture myocardial disease-state specific expression profiles in patients with myocardial infarction or heart failure with reduced ejection fraction. **a.** Schematic detailing isolation and analysis of cardiovesicles from cardiovascular cohorts (90 samples), involving patients with heart failure (HF, 15), Type I myocardial infarction (MI, 15), and control (Ctrl, 15) using the ExoCapture-MSB method. Following isolation, RNA cargo from these vesicles were sequenced. Analysis included mapping of transcripts in cardiovesicles to single cell/nuclear atlases, and differential gene expression to identify targets significantly dysregulated in HF and MI compared to Control. **b.** Venn diagram illustrating a 99% overlap among all POPDC2+ and CHRNE+ cardiovesicle transcripts across the three groups (Ctrl, HF, and MI), including 169 heart-enriched transcripts with tau score >0.6 from GTEx. Venn diagrams of the top 500 enriched transcripts (by the highest mean expression for each group) for **c.** POPDC2+ and **d.** CHRNE+ cardiovesicles in HF and MI relative to control demonstrating shared as well as unique transcripts between the cohorts. Top 25 enriched transcripts (mean expression threefold greater than standard deviation) from POPDC2+ Ctrl, HF, and MI patients were mapped onto the multiorgan single-cell transcriptomic atlas dataset (Tabula Sapiens) and a single-nuclear dataset of human control (no cardiovascular disease), dilated cardiomyopathy. Cardiovesicle transcripts from control patients mapped to the multi-organ atlas is shown as a target UMAP in **e,** with summary dot-plots and individual target UMAPs from the heart single nuclear dataset shown in **f** and **g** respectively. The same representations for the cardiovesicles from HF are shown in **h**, **i**, and **j**, while those from MI are depicted in **k**, **l** and **m**. Volcano plots displaying differentially expressed transcripts (with log FC + 1 and p < 0.01) in POPDC2^+^ cardiovesicles from patients with HF compared to control patients (**n**) with pathway enrichment analysis (**o**). Validation in the MGH validation cohort (n=30) for selected transcripts differentially expressed in HF-POPDC2^+^ EVs (MYL12B, p=0.0091, CERNA3, p=0.0178) using qRT-PCR normalized to internal and spike-in controls. Similar analysis including volcano plot for differentially expressed transcripts in POPDC2^+^ EVs in patients with MI compared to controls (**q**), pathway enrichment analysis (**r**), and experimentally validation of select transcripts in the MGH validation cohort (CDKN1A, p=0.0019, DM1-AS, p=0.0025, n= 30. Significance levels are marked as * p < 0.01, ** p < 0.001, *** p < 0.0001, calculated using the Kruskal-Wallis test.

Next, to investigate the cellular source of the cardiovesicle transcripts in disease, we identified the top 25 enriched transcripts based on their mean expression in POPDC2^+^ and CHRNE^+^ cardiovesicles in each of the three conditions separately (HF, MI, and non-cardiovascular disease control). These transcripts were subsequently mapped onto two single-cell transcriptomic atlases: an organism-wide atlas (Tabula Sapiens^29^) and a myocardial tissue atlas^24^. Control patients without cardiovascular disease exhibited cardiovesicle transcripts localizing to atrial and ventricular cardiomyocytes in the organism-wide atlas (including highly expressed cardiac-specific genes like *RYR2*, *TACC2*, *MYH7*) and predominantly featured cardiomyocyte-enriched genes in the myocardial atlas (**Fig. 5 e-g, Fig. S5 a-c**). On the other hand, while transcripts from cardiovesicles from HF and MI patients predominantly mapped to cardiomyocytes, we noted that some of the transcripts in the disease states mapped to more diverse tissues/cell types. This was broadly consistent with a well-known “metaplastic” activation of disease-related gene programs in cardiomyocytes during disease pathogenesis^30,31^. For example, in HF, organism-wide mapping suggested cardiovesicle transcripts mapped not only to cardiomyocytes, but also to endothelial cells, pulmonary cells, and skeletal muscle cells (**Fig. 5h-I, Fig. S5 d-f**). Similarly, transcripts from cardiovesicles from patients with MI were expressed in the spleen and bone marrow, alongside cardiomyocyte mapping (**Fig. 5k, Fig. S5g**), consistent with a broadly inflammatory phenotype. Analogously, mapping to our myocardial atlas revealed the activity of cardiovesicle transcripts in immune cells, endothelial cells, and fibroblasts (**Fig. 5I, j, l, m** and **Fig. S5e, f, h, i**). These data suggested the activation of pathways of inflammation and fibrosis (typically expressed in cell types other than cardiomyocytes at baseline) in cardiomyocytes themselves during disease pathogenesis.

Given the diverse cellular origin of cardiovesicular transcripts, we next studied whether differential expression <u>across</u> normal and cardiovascular disease states would reveal biological pathways relevant to myocardial tissue in these conditions. One hundred and four genes were differentially expressed in POPDC2^+^ cardiovesicles between controls and cardiovascular patients (44 for the MI group; 36 upregulated, 8 downregulated, and 60 for the HF group; 36 upregulated, 24 downregulated; p<0.05 and log FC ± 1) (**Fig. 5n, q**) while 364 genes were differentially expressed in the CHRNE^+^ cardiovesicles between controls and cardiovascular patients (86 for MI, 278 for HF; p<0.05) (**Fig. S5k, n**). The differentially expressed genes within POPDC2^+^ cardiovesicles were components of metabolic pathways such as glycolysis, and metabolic reprogramming as well as pathways associated with smooth muscle cell dysfunction, arterial hypertension, and cell adhesion molecules (CDH2 and NCAM1) (**Fig. 5o**) consistent with known dysregulation of these pathways in HF^32–37^. In contrast, genes differentially expressed in MI patient cardiovesicles showed alterations in pathways related to insulin resistance, myocardial ischemia, inflammation, peripheral tissue microangiopathy, and arterial hypertension (**Fig. 5r**). An analysis for CHRNE^+^ cardiovesicles yielded very similar results (**Fig. S5l, o**) where CHRNE^+^ cardiovesicles of HF patients contained transcripts enriched in pathways such as metabolic dysregulation and reprogramming, Glycolysis, and DNA damage response and repair (commonly dysregulated in HF^38^), whereas cargo RNAs in MI patients were components of complement-mediated inflammation, myocardial ischemia, and Frizzled signaling-related pathways consistent with their known dysregulation in cardiac ischemic disease^39–42^.

Finally, to validate the differentially expressed cardiovesicle transcripts using a complementary methodology, a set of unique genes for each disease condition was selected and prioritized (based on upset plot, coverage plots, and efficiency of functional primers, **Fig. S5 j,** see **Methods** for Target prioritization) for experimental validation in the MGH validation cohort (**Extended Table 3**). Of these, 13 targets were tested using qPCR, with 7 successfully validated after adjusting for multiple hypotheses by FDR<0.05. Among them, MYL12B and CERNA3 were significantly altered in HF vs Ctrl, while CDKN1C and DM1-AS were differentially expressed in MI vs Ctrl POPDC2^+^ cardiovesicles (**Fig. 5p, s**). For the CHRNE^+^ cardiovesicles, MT-ND1 and MT-ND5 were significantly expressed in HF vs Ctrl, and CTF1 expression was altered in MI vs Ctrl **Fig. S5 m, p**). While we only chose to validate a subset of these differentially expressed genes using complementary assays (qRT-PCR) in a separate validation cohort, our data support the robustness of the data. Collectively, these findings demonstrate that cardiovesicles not only contain cardiac-specific genes but also exhibit a disease-induced transcriptomic shift, mirroring the underlying pathophysiological changes in cardiac diseases. Their capacity to map disease-specific pathways and gene expression profiles provides a valuable non-invasive window into the cellular processes driving these conditions.

### Molecular profiling of cardiovesicles from patients with aortic stenosis reveals enrichment of transcripts expressed in cardiomyocytes and relevant pathways in myocardial remodeling

To provide evidence of the cardiomyocyte origin of circulating cardiovesicles, we extracted cardiovesicles (using POPDC2 immunocapture) from a cohort of patients with aortic stenosis (AS) and controls (VUMC Cohort, **Table S4**) who had cardiac tissue available for snRNAseq analysis (see **Methods** for details of cohort, snRNAseq also separately analyzed as part of another study) (**Fig. 6a**). We performed bulk RNA-sequencing on the input plasma EVs before cardiovesicle extraction (EV_AS) and cardiovesicles (POPDC2_AS and POPDC2_Ctr). Single-nuclear RNA sequencing (snRNA-seq) of cardiac tissue from the same patients was performed (**Fig. 6a**). By comparing the most abundant transcripts in each of these groups (top 150 by mean expression across samples in each group), we identified transcripts unique to cardiovesicles (present in both POPDC2_AS and POPDC2_Ctr, “Cardiovesicle_Unique”) and those unique to bulk EVs (“BulkEV_Unique”) (**Fig. 6b**). These transcripts were mapped to the Tabula Sapiens (TS)^29^ single-cell atlas and classified based on their relative expression in different tissues. Consistent with prior organism-wide mapping studies, BulkEV_Unique transcripts were widely sourced from organs across the entire body (notably from hematopoietic and blood cells), whereas Cardiovesicle_Unique transcript origin was more cardiac-specific (**Fig. 6c-f**). We then mapped the BulkEV_Unique and Cardiovesicle_Unique transcripts from each AS patient to their corresponding cardiomyocyte expressions derived from tissue snRNA-seq of the same individual. Our analysis across eleven AS patients demonstrated that the cardiovesicle extraction process selectively enriches for genes predominantly expressed in cardiomyocytes (**Fig. 6g, h**). This pattern persisted both cumulatively, when selecting the top 150 enriched genes from all patients, and individually, when selecting top enriched genes for each individual patient, indicating personalized enrichment with each set of top enriched genes being both cardiac-specific and unique to each patient within the POPDC2^+^ cardiovesicles (**Fig. S6 a-b)**. Notably, despite significant heterogeneity of expression of transcripts across individual patients (as is known in these patients with aortic stenosis), the top cardiovesicle transcripts demonstrated consistent mapping to cardiomyocytes from the same patient.

**Figure 6.**
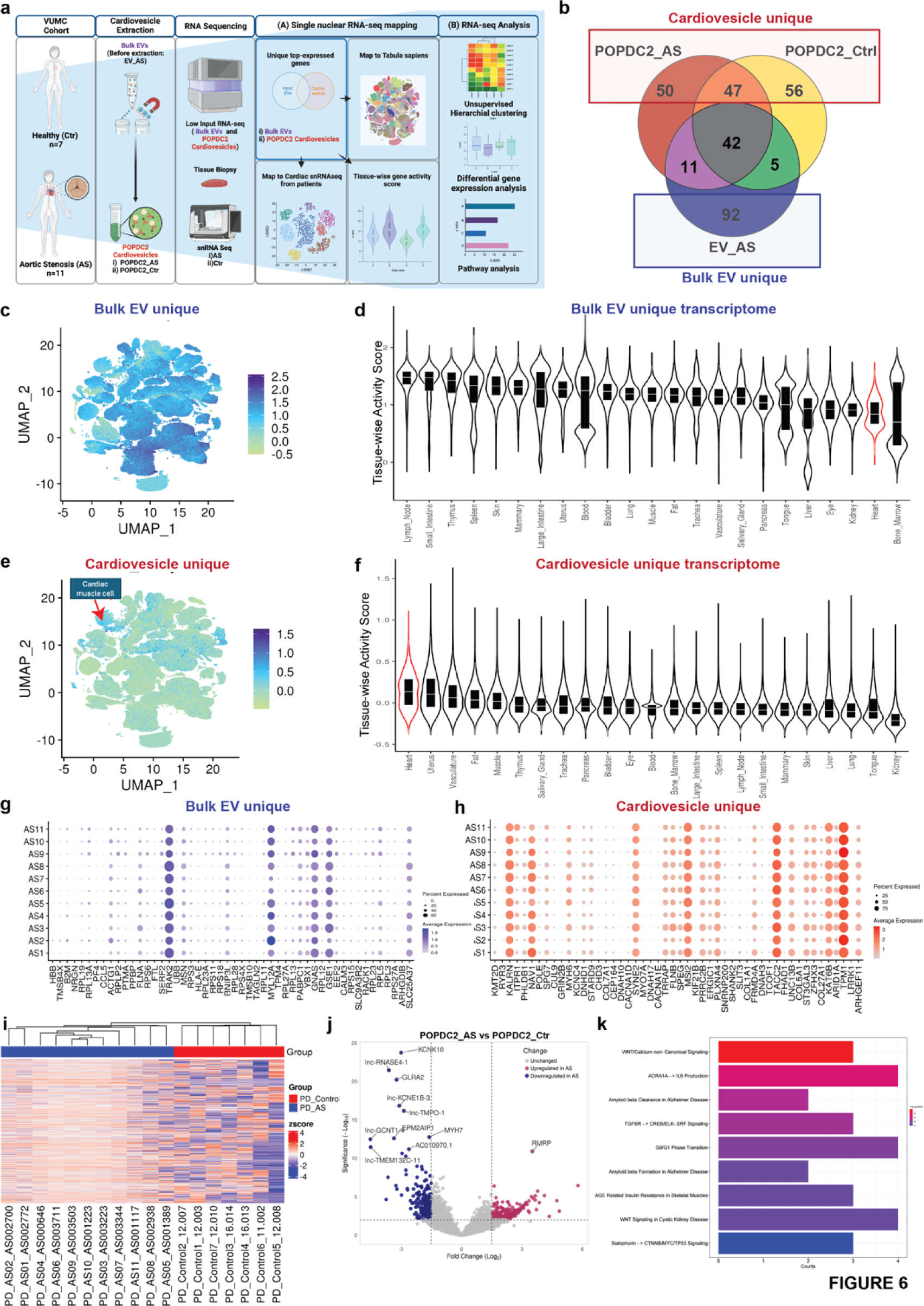
Molecular profiling of cardiovesicles from patients with aortic stenosis reveals enrichment of transcripts expressed in cardiomyocytes and relevant pathways in myocardial remodeling. **a.** Schematic overview detailing the isolation and transcriptomic analysis of cardiovesicles from the AS cardiovascular cohort (18 samples), including Bulk EVs (unprocessed EVs, no cardiovesicle extraction) from AS patients (EV_AS, n=11) and POPDC2+ cardiovesicles from AS patients (POPDC2_AS, n=11) and controls (POPDC2_Ctr, n=7). Analysis of the top abundant transcripts, prioritized by mean expression, of EV_AS, POPDC2_AS, and POPDC2_Ctr, was conducted to identify genes unique to EV_AS (labeled as ‘Bulk EV_unique’) and those unique to POPDC2+ cardiovesicles (POPDC2_AS and POPDC2_Ctr; labeled as Cardiovesicle_unique). These were mapped onto single nuclear RNA-seq datasets and differential expression analysis was performed to identify and validate targets significantly dysregulated in AS compared to Control. **b.** Venn diagram analysis of the top 150 abundant transcripts (prioritized by mean expression) of EV_AS, POPDC2_AS, and POPDC2_Ctr showing Bulk EV_unique and Cardiovesicle_unique transcripts. Distribution of Bulk EV_Unique and (**c**) and cardiovesicle_unique transcripts (**e**) represented as UMAPs of the multiorgan single cell transcriptomic atlas dataset (Tabula Sapiens), with their respective tissue-wise transcriptional activity score showing other tissues in Bulk EV_unique (**d**) and heart in Cardiovesicle_unique gene sets (**f**). Dot plots **g.** and **h.** show the expression counts of Bulk EV_unique and Cardiovesicle_unique transcripts in cardiomyocytes of each individual AS patient, respectively. **i.** Unsupervised hierarchical clustering illustrates the transcriptional differences between AS (POPDC2_AS) and Ctrl (POPDC2_Ctr) POPDC2+ cardiovesicles**. j.** Volcano plots represent differentially expressed genes between POPDC2_AS and POPDC2_Ctr **k.** Pathway enrichment analysis reveals the differentially expressed genes between AS and Ctr POPDC2+ cardiovesicles. log FC cutoff +1, and FDR <1% have been used as statistical significance cut-offs for the differential gene expression analysis

Finally, we sought to examine the transcriptional differences between the cardiovesicles of AS patients (POPDC2_AS) and controls (POPDC2_Ctr). Unsupervised hierarchical clustering based on gene expression patterns revealed distinct separation between the POPDC2_AS and POPDC2_Ctr groups, with unique expression profiles within the cardiovesicles of each group (**Fig. 6i**). Differential gene expression analysis (DGEA) between the two groups (with FDR cutoff <0.05) identified 578 DEGs, comprising 334 protein-coding (103 upregulated and 231 downregulated) and 244 non-coding genes (55 upregulated and 189 downregulated), as indicated in the volcano plot (**Fig. 6j**). Pathway enrichment analysis on all DEGs between POPDC2_AS and POPDC2_Ctr identified significant involvement of several pathways, including the Wnt signaling pathway, inflammation mediated by chemokine and cytokine signaling, amyloid response, and TGF-beta signaling, all of which are implicated in cell proliferation, inflammatory processes, protein deposition, and fibrosis characteristic of AS^43–47^ (**Fig. 6k**). In summary, our comprehensive analysis confirms that cardiovesicles isolated from patients with AS are enriched with cardiomyocyte-specific transcripts and reveals distinct transcriptional profiles between AS patients and controls, highlighting the potential of cardiovesicles as a source for understanding disease mechanisms and identifying potentially novel biomarkers for CVD such as aortic stenosis.

## DISCUSSION

The search for a circulating “liquid” surrogate for human tissue states is a prime objective of precision personalized medicine. Oncology has been at the vanguard of this goal, realizing the clinical potential of liquid biopsy in clinical management via leveraging the unique genomic and transcriptional instability of cancer to differentiate normal from disease (e.g., cell-free DNA^48–51^, EV^52–54)^. While plasma molecular profiling studies in equally debilitating conditions such as cardiovascular diseases have offered some insights into broad mechanisms^55,56^, direct, dynamic tissue phenotyping of the heart has remained elusive due to ethical challenges, the unfavorable risk-benefit ratio of myocardial biopsies, and the limitations in cardiac specificity of plasma biomarkers. Here, we introduce a novel paradigm to isolate and characterize subpopulations of EVs—small circulating vesicles bearing molecular cargo characteristic of their source cell—enriched from a cardiomyocyte source in human circulation. Through an integrated, multi-parametric design that leveraged advanced *in vitro*, *in silico*, mouse, and clinical sampling, we identified two protein markers on the surface of cardiomyocyte-derived EVs (cardiovesicles)—POPDC2 and CHRNE—that facilitated immunocapture of cardiovesicles in cellular, murine, and human systems, providing a quantitative, plasma-based sample of the cardiomyocyte transcriptome. Importantly, POPDC2^+^ and CHRNE^+^ EVs reflect the cardiomyocyte transcriptome in a dynamic fashion, mapping to health and disease states across the spectrum of ischemic CVD to heart failure. These results suggest a utility for cardiovesicles in assessing myocardial health and pathology in a serial, non-invasive fashion across a wide array of cardiovascular disease states and provide a roadmap for analogous strategies across human biology.

The search for tissue specificity within circulating biomarker space is an emerging space. Recent work assigning tissue specificity to the circulating proteome via tissue gene expression demonstrated a striking relation of brain- and cardiac-enriched proteins to organ-relevant clinical phenotypes^9^. However, direct concordance between circulating molecular biomarkers (proteins or whole blood transcripts) and tissue molecular states are confounded by several key factors, most prominent of which include (1) poor protein-to-RNA concordance^57^ and (2) impact of unique patient-level factors that impact transcription that limit the ability to “deconvolute” tissue specificity of a circulating biomarker across individuals. In turn, leading cardiac-enriched candidate proteins from transcriptional prioritization included clinically recognizable biomarkers that reflect end-phenotypes of CVD (troponin [TNNT2]^58^, B-type natriuretic peptide [NPPB]^11^), not upstream markers of intervenable physiology and dysfunction. State-of-the-art methods for transcriptional deconvolution (e.g., transfer learning) have attempted to address these challenges, though many still use normative references or are confined to deconvolution of cell type from tissue bulk RNA-seq^59–61^, limiting their generalizability.

Here, we overcome these obstacles by isolating cardiomyocyte-specific EVs (coined “cardiovesicles”) from an individual to sample that individual’s cardiac state uniquely. Through a comprehensive multi-dimensional translational approach integrating mass spectrometry and deep learning membrane protein prediction, we screened ≈4-5K proteins across iPSC-cardiomyocytes and their derived EVs with subsequent filtering for cardiac specificity (at both protein and transcriptional level in human cardiac tissue samples [GTEx]) and antibody performance, arriving at two candidates, POPDC2 and CHRNE. Both targets were physiologically plausible as cardiomyocyte-specific targets: POPDC2 (popeye domain containing 2) is highly expressed in cardiomyocyte membranes and implicated in regulation of cardiac conduction^62,63^, and CHRNE (nicotinic acetylcholine receptor epsilon) is expressed both on cardiomyocytes and skeletal muscle and may be involved in muscular end-plate neurotransmission^64,65^. POPDC2 and CHRNE were shown to decorate the surface of iPSC cardiomyocyte-derived EVs via state-of-the-art single EV imaging techniques and contained a broad array of overlapping cardiac-enriched transcripts implicating known cardiomyocyte metabolic pathways, supporting selectivity. As opposed to “bulk” EVs of diverse organ origin, across genetically engineered models systems (ExoMap1), *in vitro* iPSC models, and human plasma, human cardiovesicles contained cardiomyocyte-derived transcripts, specifically dynamic in disease conditions: individuals with MI or HF harbored transcripts across a broad array of well-known, inducible pathways within cardiomyocyte under stress (inflammation, fibrosis, structural and metabolic remodeling, among them). Strikingly, pathways highlighted by cardiovesicle transcripts appeared to be distinct <u>between</u> MI and HF and reflected predominantly pro-inflammatory ischemic pathogenesis for MI and metabolic pathogenesis for HF, consistent with the known <u>tissue</u> pathobiology in these states^66–68^. Importantly, this approach yielded a more expansive set of biomolecules more specific for ischemic and HF pathobiology than standard clinical biomarkers like troponin or natriuretic peptides (MYL12B, CERNA3, CDKN1C, and DM1-AS), including known <u>functional</u> epigenetic markers for cardiac remodeling (e.g., miR-30d^27^). Finally, in participants with concurrent plasma and myocardial sampling, we found that transcripts unique to cardiovesicles (and not highly expressed in bulk EVs) largely exhibited cardiac selectivity in whole organism transcriptional atlases (Tabula sapiens^29^) and mapped to cardiomyocytes in single nuclear myocardial RNA-seq from the same individuals.

These “bench-to-bedside” results specify POPDC2 and CHRNE as candidate markers for circulating cardiomyocyte EV “liquid biopsy” and showcase the utility of this paradigm more generally as a discovery platform for other organ systems. Our results come in the context of an emerging hunt for a cardiac-specific EV population in circulation. For example, several studies have suggested CD172 as a cardiac-specific marker, including as a marker of cardiomyocyte-specific EVs dysregulated in diseases such as aortic stenosis^69,70^. Nevertheless, CD172 appears to have a more diverse tissue expression beyond cardiomyocytes with copious expression in the brain, immune cells and kidney (Human Protein Atlas^71^). The use of a unique genetically engineered cardiac-specific EV tracking murine model system (ExoMap1^25^) for trans-species replication increased confidence in our results. After implicating POPDC2 and CHRNE as EV surface markers in human iPSC models, GTEx^72^, and snRNA-seq, we confirmed their presence on EVs of definite cardiomyocyte origin in the murine model (and their notable absence on immune-related EVs in the same model). The implications of this bidirectional approach are significant, enabling evaluation of cardiomyocyte-relevant “liquid biopsy” across human disease states, populations, and in model systems <u>dynamically</u> and <u>serially</u>.

The ability to detect cardiomyocyte-specific states in human circulation is a major boon to human translational precision medicine in CVD, with applications to phenotype early CVD predisposition (potentially before canonical biomarkers are elevated) to examining heterogeneity in CVD states in more advanced disease that may inform mechanism and therapy (e.g., HF).

Nevertheless, several important limitations merit mention. As with any immunocapture technique, differences in EV yield depend on antibody efficiency and variability. While our results were consistent across models and human tissues, we envision broader clinical translation will require more process optimization (e.g. monoclonal antibodies or microfluidics^73,74^). In addition, POPDC2^+^/CHRNE^+^ cardiovesicles likely reflect a subpopulation of cardiac-derived EVs (given our prioritization for specificity for cardiomyocyte expression). While RNA cargo of these cardiovesicle subpopulations likely do not reflect the <u>entire</u> intracellular cardiomyocyte transcriptome, our results indicate that the transcripts sampled within these cardiovesicles reflect disease-specific pathway alterations characteristic of clinical pathogenesis within the source cells. Of note, even canonical EV markers (e.g., CD63/CD81/CD9) are known to mark distinct subpopulations of EVs with distinct cargo^75^. Most of the RNAs within these cardiovesicles are likely fragments (as determined by the 5’ to 3’ coverage plots of the RNA sequencing results), we did not endeavor to determine if some may be full-length. Furthermore, our approach here interrogated primarily RNA cargo within EVs, and future studies will broaden the scope of the cardiovesicle cargo to other epigenetic and protein-based mediators of CVD pathogenesis. Ultimately, we envision this cardiomyocyte-focused approach as a paradigm for discovery, complementing ongoing research into other cell types^76^. Future work will focus on optimizing these parameters alongside advances in sequencing technology, low-input library construction methods, and antibody standardization.

In conclusion, using high-throughput mass spectrometry, genetic and human models of EV biology, and patient samples, we identified and characterized a unique subset of EVs within human circulation that are cardiomyocyte-derived. Transcripts within these “cardiovesicles” were enriched in human cardiomyocytes, reflected pathways of human myocardial dysfunction, and were distinct across health and disease. Importantly, these findings did not rely solely on known published reference profiles, instead reflecting a patient-specific cardiomyocyte transcriptional profile. These results provide a foundation for transitioning modern molecular discovery in CVD from circulation in human populations toward a tissue-enriched paradigm, a credible “liquid biopsy” of the human cardiomyocyte. Expanding on this approach across specific cell types, organs, and serially within different types of CVD may significantly enhance patient-specific subphenotyping, targeting, and therapy, opening new avenues in precision medicine for understanding the effector functions within cardiomyocytes in a non-invasive fashion.

## Data Availability

All data produced in the present study are available upon reasonable request to the authors

## Supplementary Table and Data Figures

**Table S1b: Proteomics discovery data**

**Table S2:**
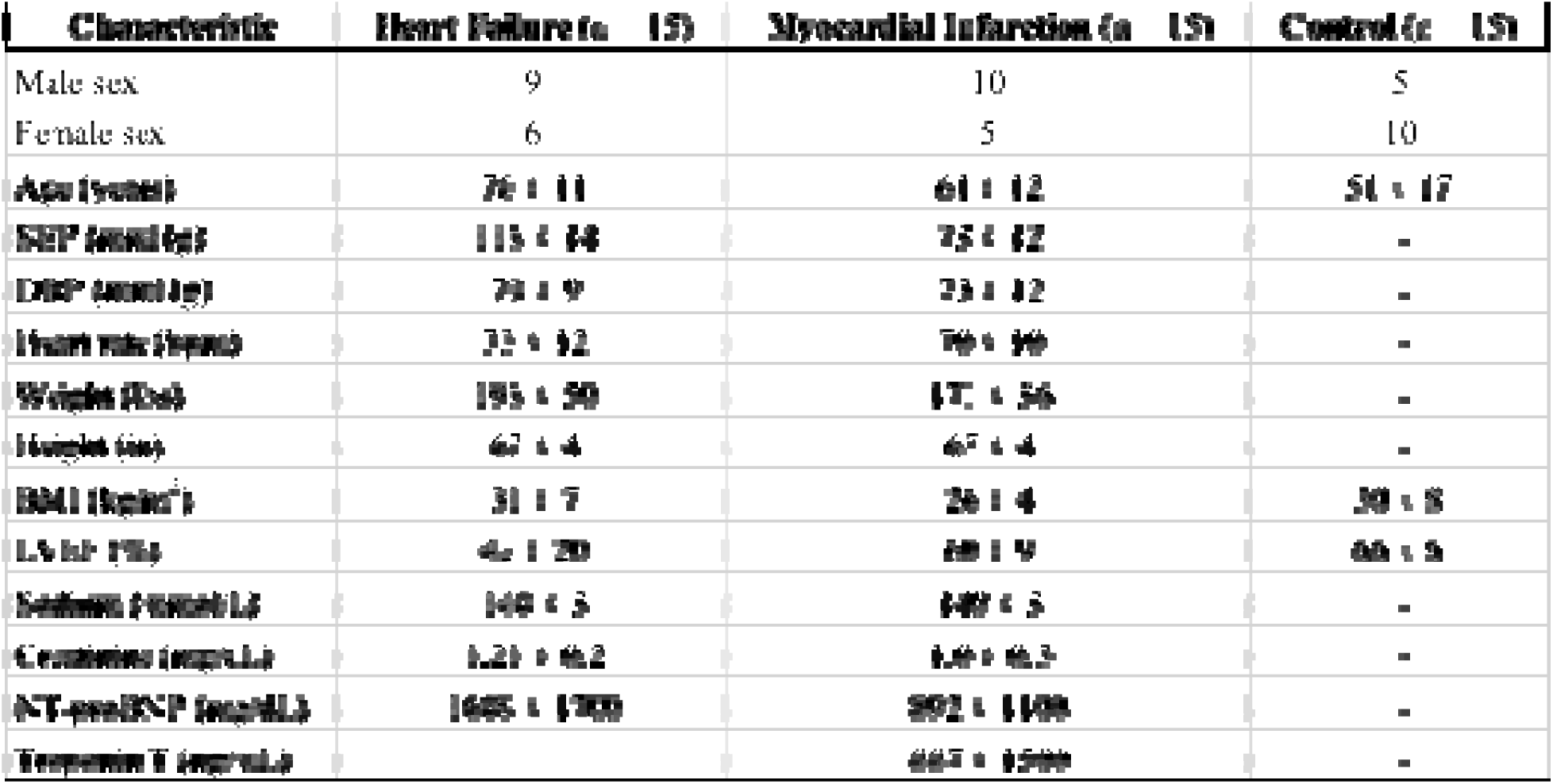
MGH Cohort Discovery-Clinical demographics.

**Table S3:** MGH Cohort- validation- Clinical demographics.

| Characteristic | Heart Failure (n = 15) | Myocardial Infarction (n = 15) | Control (n = 15) |
| --- | --- | --- | --- |
| Male sex | 7 | 7 | 7 |
| Female sex | 8 | 8 | 8 |
| Age (years) | 65 ± 10 | 64 ± 11 | 64.5 ± 10.5 |
| SBP (mmHg) | 118 ± 14 | 122 ± 15 | - |
| DBP (mmHg) | 72 ± 9 | 74 ± 12 | - |
| Heart rate (bpm) | 75 ± 12 | 72 ± 10 | - |
| Weight (lbs) | 195 ± 50 | 170 ± 36 | - |
| Height (in) | 67 ± 4 | 67 ± 4 | - |
| BMI (kg/m<sup>2</sup>) | 31.0 ± 7 | 26.5 ± 4 | 30.4 ± 8 |
| LVEF (%) | 55 ± 12 | 57 ± 10 | 66 ± 9 |
| Sodium (mmol/L) | 140 ± 3 | 140 ± 3 | - |
| Creatinine (mg/dL) | 1.22 ± 0.2 | 1.13 ± 0.3 | - |
| NT-proBNP (pg/mL) | 1500 ± 1600 | 890 ± 1100 | - |
| Troponin T (ng/mL) | 6 ± 12 | 385 ± 1490 | - |

**Table S4:** Clinical demographics of donors and individuals with aortic stenosis (single-nuclear RNA-sequencing)

| **Characteristic** | **Aortic Stenosis (n=11)** | **Control (n=7)** |
| --- | --- | --- |
| Age, years | 65.6 ± 19.3 | 47.6 ± 15.5 |
| Gender, Male, % | 55 (6/11) | 57 (4/7) |
| Coronary artery disease | 73 (8/11) | - |
| History of heart failure | 100 (11/11) | - |
| Hypertension, % | 91 (10/11) | - |
| Hyperlipidemia, % | 70 (7/10) | - |
| Diabetes, % | 73 (8/11) | - |

**Figure S1.**
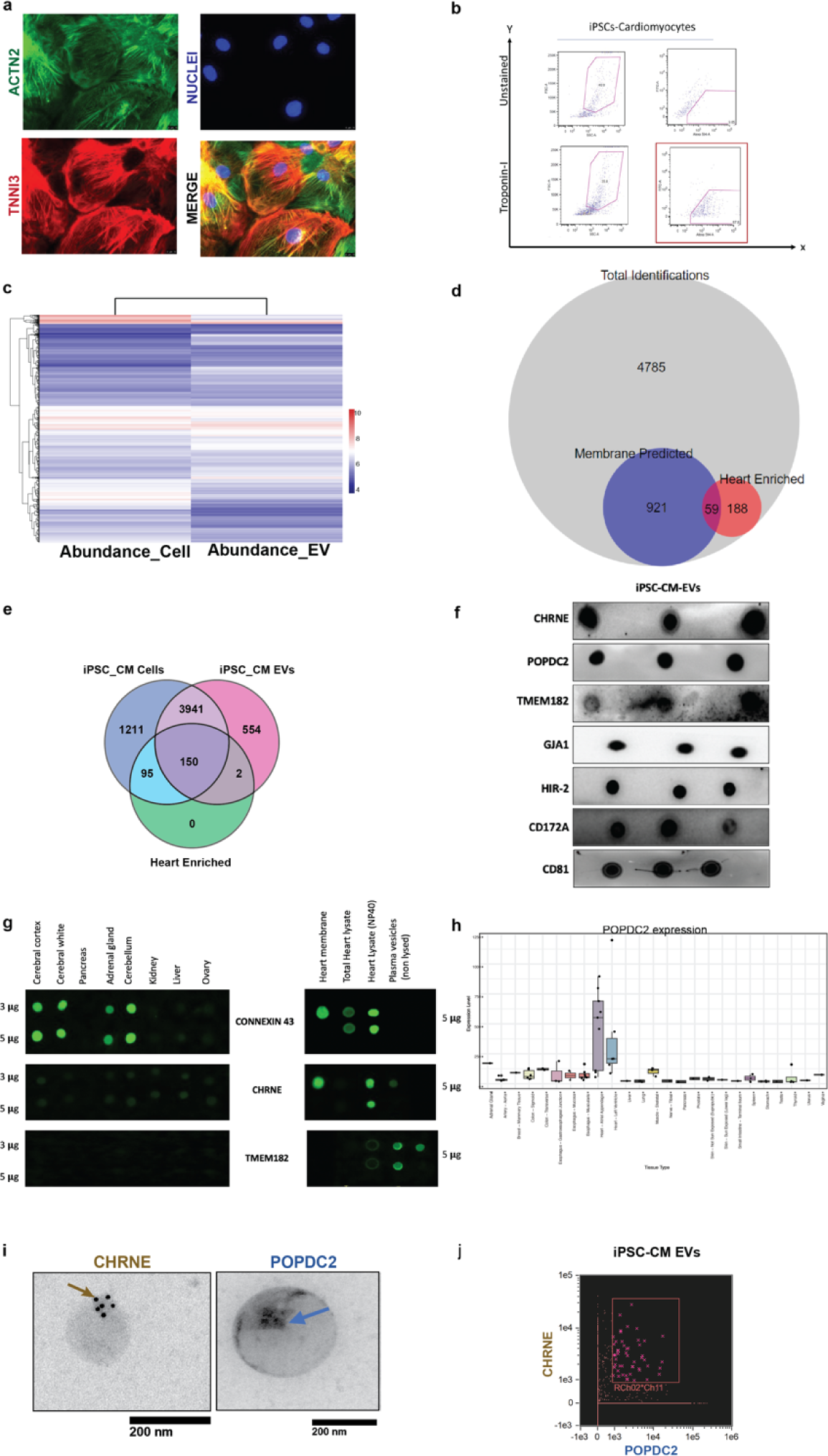
Discovery and experimental validation to identify POPDC2 and CHRNE as cardiomyocyte-EV (Extracellular Vesicle) membrane protein candidates. Representative **a.** confocal images (showing ACTN2 and TNNI3 positivity) and **b.** Flow-cytometric data (showing Troponin-1 expression) of iPSC-CM (induced Pluripotent Stem Cell-derived Cardiomyocytes) demonstrating cardiac specificity by their expression of classical cardiac markers such as ACTN2, TNNI3 and TNNT1. **c.** Heatmap showing the abundance of proteins expressed in iPSC-CMs and iPSC-CM-EVs (iPSC-CM Extracellular vesicles) as identified using LC-MS/MS (Liquid Chromatography with tandem mass spectrometry). **d.** Proportional venn diagram showing the intersection of total proteins identified using LC-MS/MS, membrane -localize and heart enriched. **e.** Venn diagram showing 150 targets that were identified using LC-MS/MS present in iPSC-CM, iPSC-CM-EVs and enriched in the heart (from Human Protein Atlas data). **f.** Representative experimentally validated dot blots in iPSC-CMs for various antibodies against proteins identified using the proteomic approach discovery. **g.** Representative tissue-specific dot blots using brain and heart tissue lysates from human samples tested for CONNEXIN43, TMEM182 and CHRNE derived from Computational approach. **h.** Tissue-wise proteomic expression of POPDC2 using GTEx showing elevated expression in heart tissue. **i.** Representative images of POPDC2^+^ (blue arrows) and CHRNE^+^ (gold arrows) EVs in iPSC-CM-EVs showing single positive (POPDC2^+^ CHRNE^-^; POPDC2^-^ CHRNE^+^) EVs visualized through Transmission Electron Microscopy with gold particle POPDC2 (5 mm) and CHRNE (12 mm). **j.** POPDC2^+^ and CHRNE^+^ EVs in iPSC-CMs demonstrated using ImageStream flow cytometry based-methodology (n=3 replicates).

**Figure S2.**
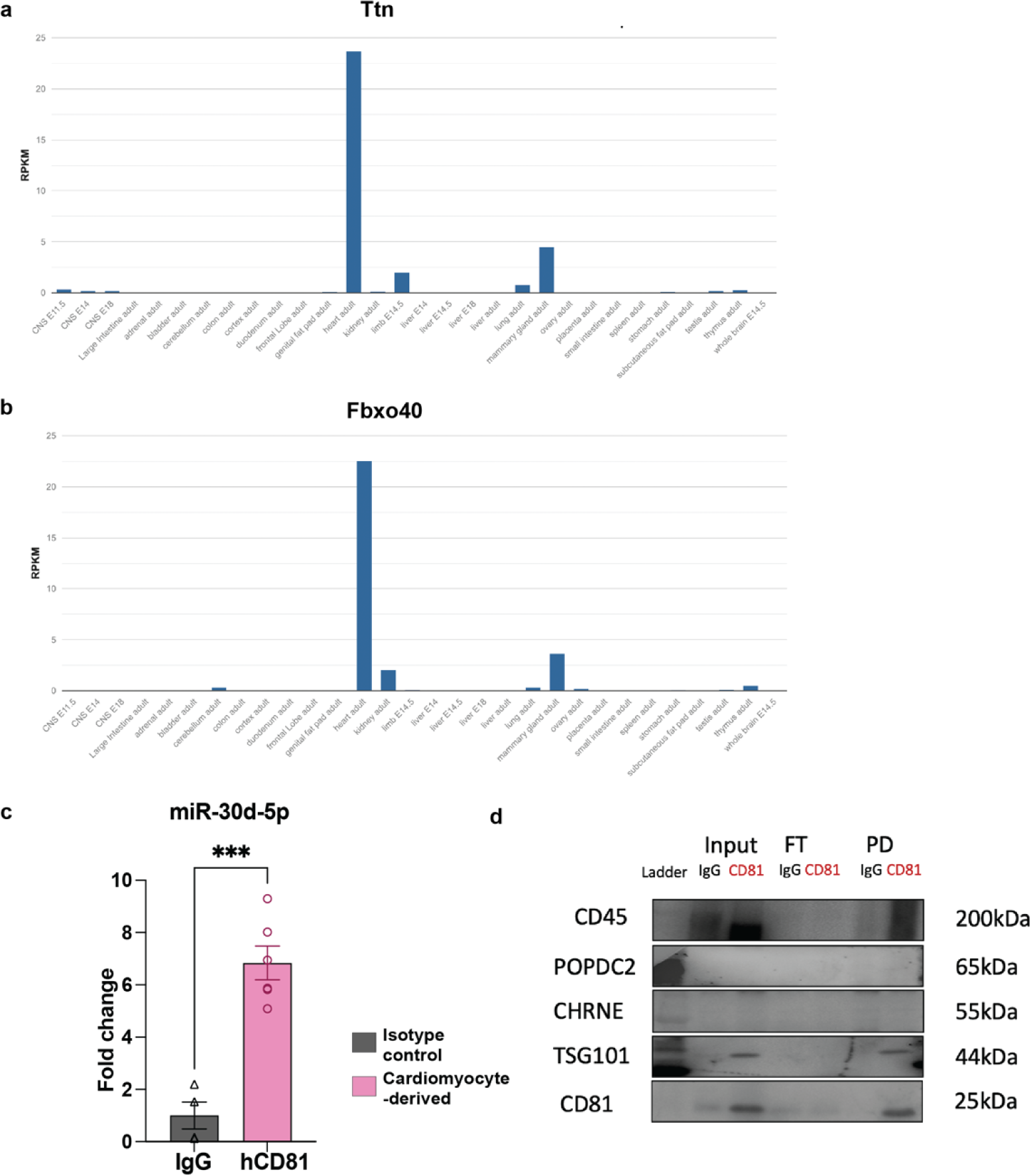
Cardiac-specific transcriptome of CD81 EVs in αMHC-Cre driven Exomap mouse model. **a,b.** Representative tissue-wise expression of heart-enriched transcripts (Ttn, Fbxo40) obtained using Mouse ENCODE transcriptomic data from NCBI. **c.** Enrichment of cardiac-specific small RNA miR-30d -5p in immunocaptured cardiac-enriched humanized CD81 EVs compared to the respective isotype control. **d.** Immune tissue-specific Exomap mouse model does not express POPDC2 and CHRNE as seen using the western blot image showing isolated hsCD81+ EVs (enrichment for CD81 expression in pulled-down samples as well as other EV markers such as Alix and TSG101) from the plasma of mice derived from crossing vav-cre transgenic mice (cre expression in hematopoietic cells ) and ExoMap mice. These EVs were enriched for immune markers (CD45), but not POPDC2 or CHRNE.

**Figure S3.**
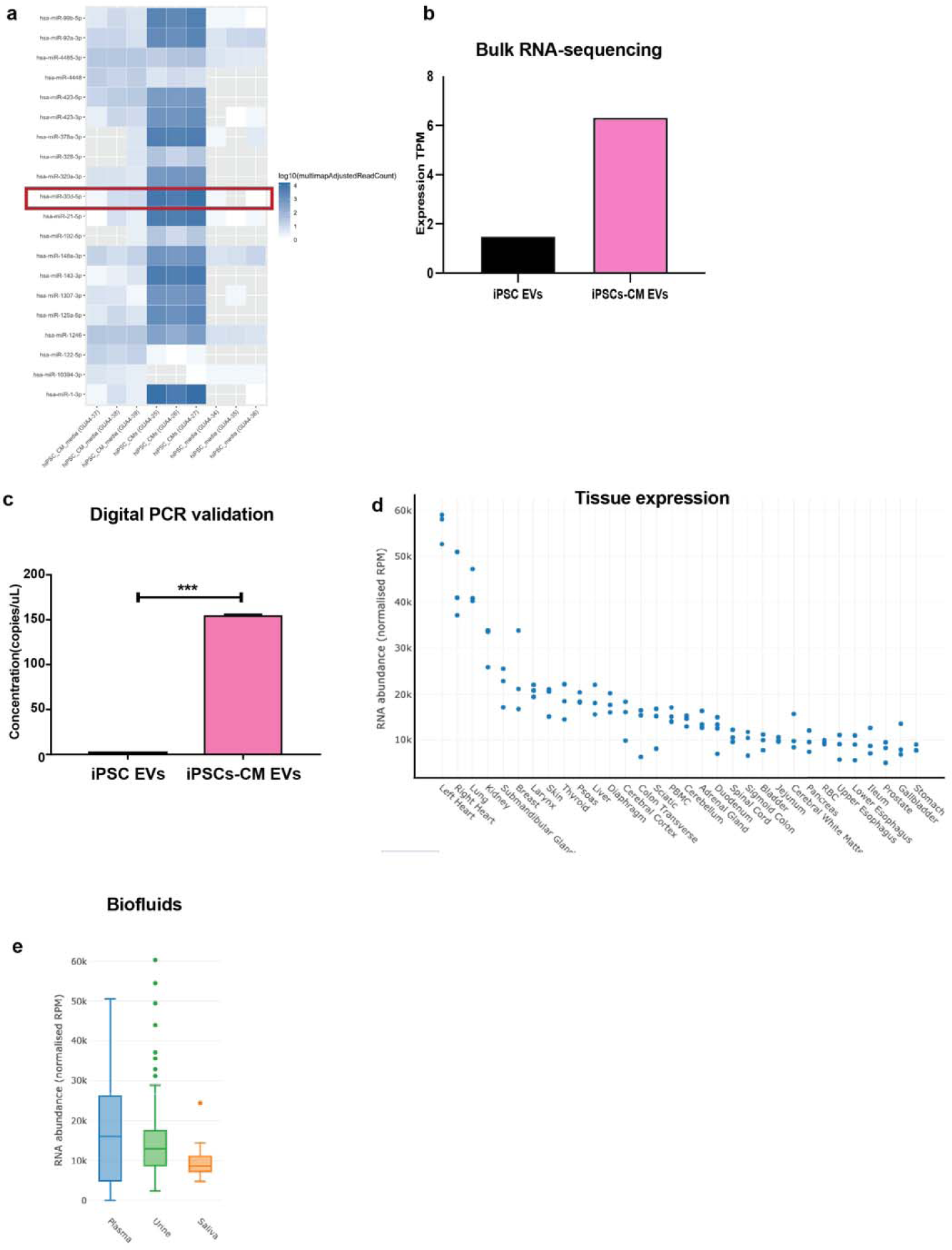
Small RNA sequencing of iPSC-CMs (induced Pluripotent Stem Cell-derived cardiomyocytes) and iPSC-CM-EVs (induced Pluripotent Stem Cell-derived cardiomyocyte – Extracellular Vesicles). **a**. Heatmap of the most significantly different miRNAs between iPSC-CMs and iPSC-CM-EVs with human Pluripotent Stem Cells as controls with miRNA-30d (miR-30d) as one of the most enriched in iPSC-CMs and iPSC-CM-EVs. **b.** Expression of miR-30d as observed in bulk RNA-sequencing showing enrichment in iPSC-CM-EVs compared to iPSC-CMs. **c.** Validation of miR30d expression using digital PCR showing enrichment in iPSC-CM-EVs compared to iPSC-CMs (p-value calculated using student t-test). **d.** Expression of miR-30d in different human tissues showing cardiac enrichment. **e.** Expression of miR-30d in different human biofluids showing expression in human plasma.

**Figure S4.**
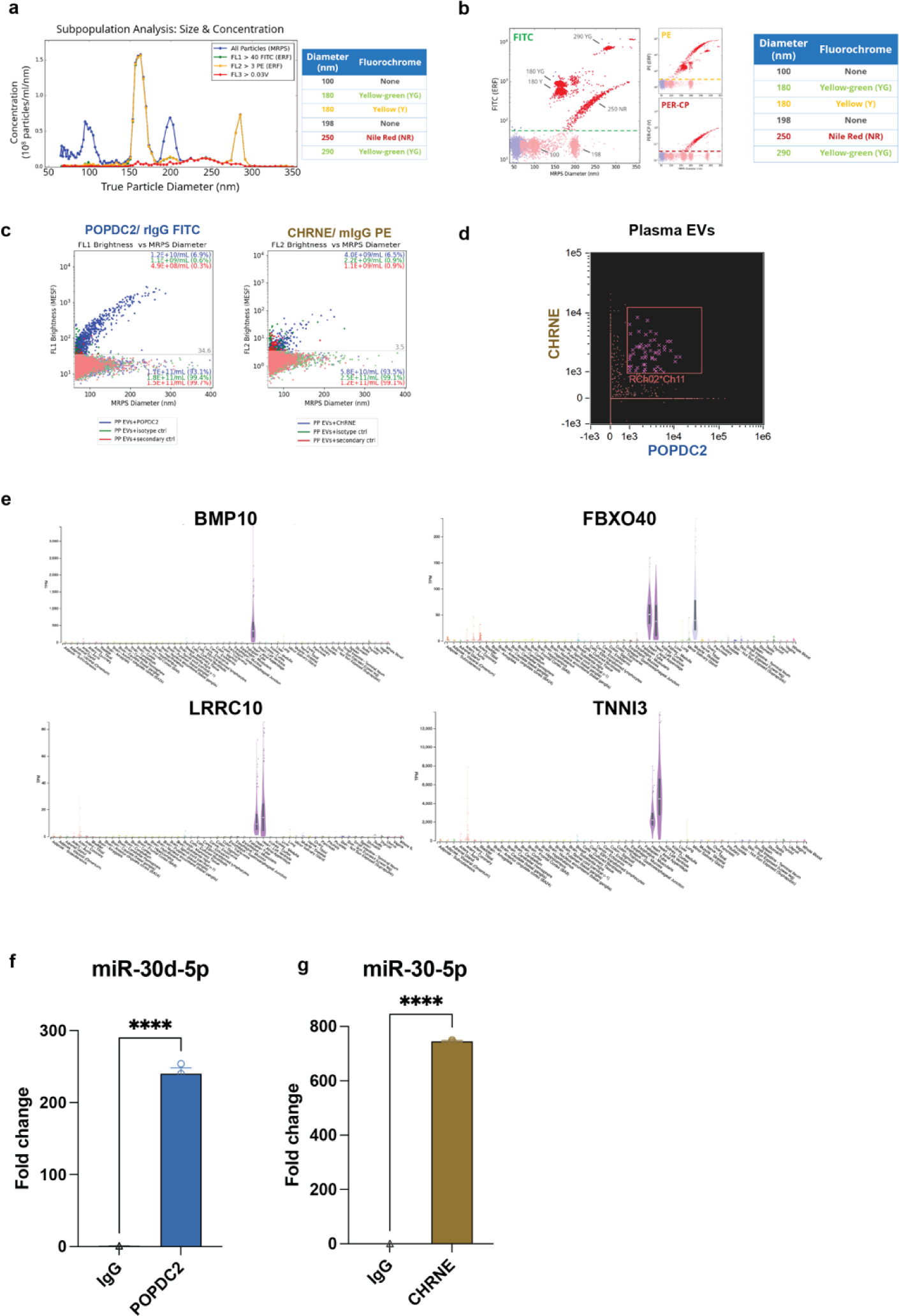
Characterization and profiling POPDC2^+^ and CHRNE^+^ cardiovesicles in healthy human plasma. Appropriate Controls for Fluorescent-Microfluidic Resistive Pulse Sensing (F-MRPS) **a. b.** demonstrating the size and fluorescence specificity using appropriate bead-controls. **c.** Isotype and secondary antibody only control for POPDC2^+^ and CHRNE^+^ immunolabelling. **d.** POPDC2^+^ and CHRNE^+^ EVs in healthy pooled plasma EVs demonstrated using ImageStream flow cytometry based-methodology. **e.** Tissue-wise transcriptomic expression of mRNA transcripts enriched in POPDC2 and CHRNE cardiovesicles (BMP10, FBXO40, LRRC10, TNNI3) using GTEx showed elevated expression in human heart tissue. Enrichment of cardiac-specific small RNA miR-30d -5p in **f.** POPDC2 and **g.** CHRNE immunocapture from healthy human plasma EVs compared to the respective isotype control.

**Figure S5.**
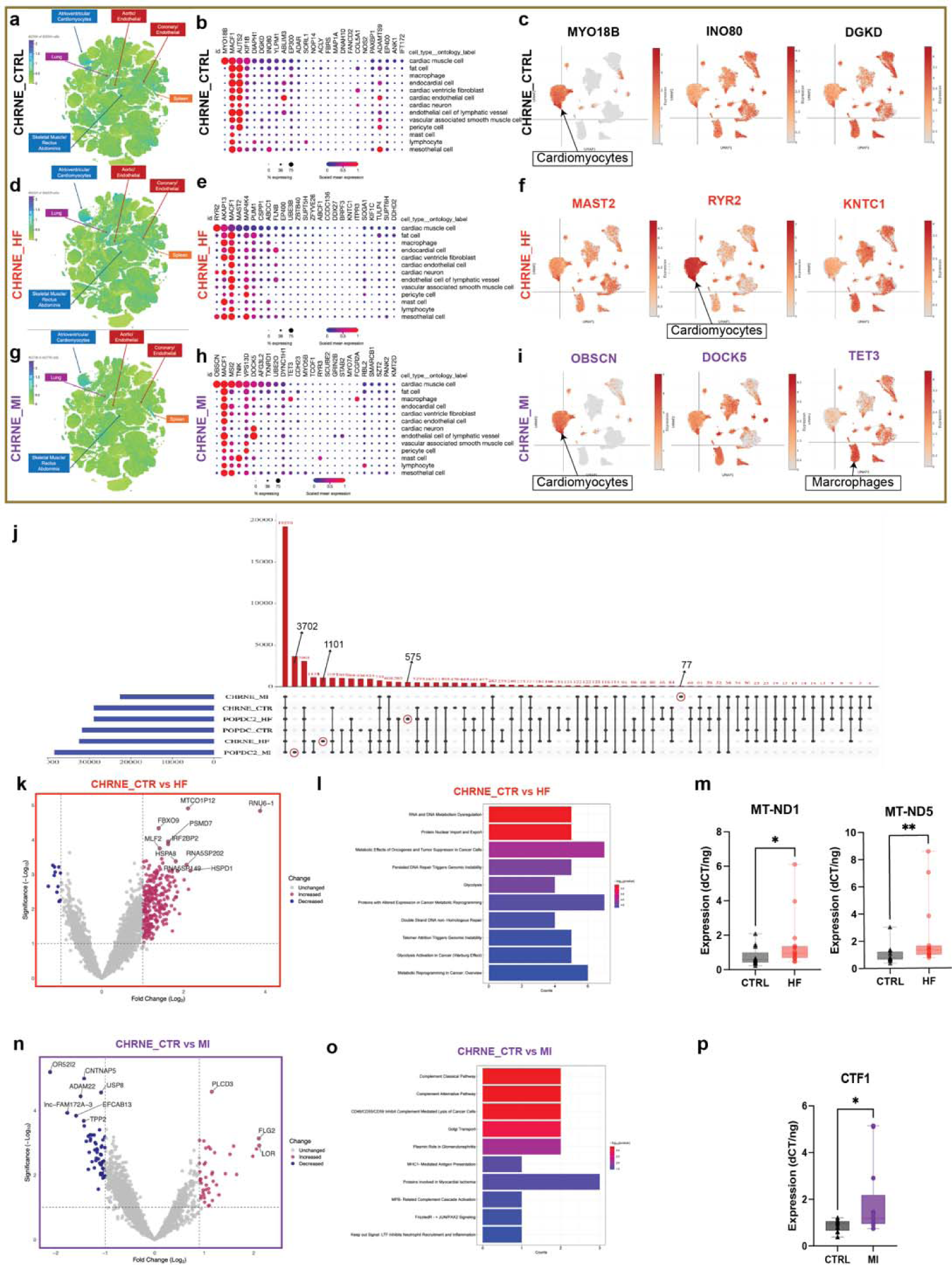
Analysis of CHRNE^+^ cardiovesicle transcriptome from HF and MI patients with respect to control patients in the MGH cardiovascular cohort. Top 25 enriched transcripts (with mean expression threefold greater than standard deviation) from CHRNE^+^ Ctrl, HF and MI patients were identified and mapped onto the multiorgan single cell transcriptomic atlas dataset (Tabula Sapiens) (**a, d, g** respectively) and a single-nuclear dataset of human control (no cardiovascular disease), dilated cardiomyopathy (**b, c, e, f, h, i).** Cardiovesicle transcripts from control patients mapped to the multi-organ atlas is shown as a target UMAP in **a,** with summary dot-plots and individual target UMAPs from the heart single nuclear dataset shown in **b** and **c** respectively. The same representations for the cardiovesicles from HF are shown in **d**, **e**, and **f**, while those from MI are depicted in **g**, **h** and **i**. **j.** Upset plot showing common and unique transcripts in cardiovesicles isolated from Ctrl, HF and MI patients using POPDC2 and CHRNE immunocapture. Genes expressed in 50% of each group of samples are shown. The unique number of genes for each group are indicated using a black arrow with the respective number of genes for that group (3702 genes unique to POPCD2 MI, 1101 unique to CHRNE HF, 575 unique to POPDC2 HF, and 77 genes unique to CHRNE MI).The unique transcripts for each group is indicated using a black arrow with the respective number of transcripts for that group. Transcripts within Cardiovesicles immunocaptured with CHRNE from Ctrls showed high cardiac specificity compared to HF and MI. Volcano plots displaying differentially expressed transcripts (with log FC + 1 and p < 0.01) in CHRNE^+^ cardiovesicles from patients with HF compared to control patients (**k**) with pathway enrichment analysis (**l**). **m.** Validation in the MGH validation cohort (n=30) for selected transcripts differentially expressed in HF-CHRNE^+^ EVs (MT-ND1, p=0.0253, MT-ND5, p=0.0061) using qRT-PCR normalized to internal and spike-in controls. Similar analysis including volcano plot for differentially expressed transcripts in CHRNE^+^ EVs in patients with MI compared to controls (**n**), pathway enrichment analysis (**o**), and experimentally validation of select transcripts in the MGH validation cohort (**p,** CTF1, p=0.0398, n= 30. Significance levels are marked as * p < 0.01, ** p < 0.001, *** p < 0.0001, calculated using the Kruskal-Wallis test.

**Figure S6.**
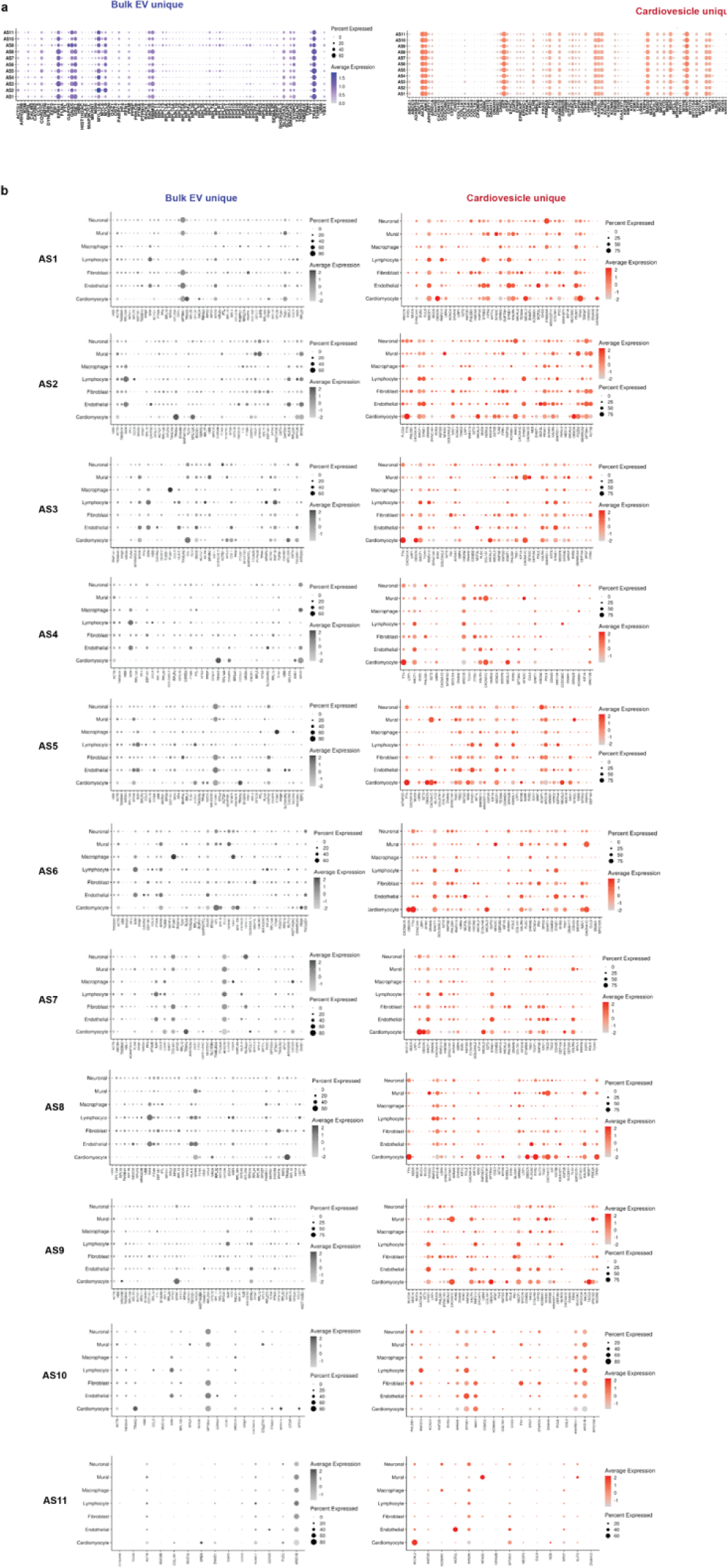
Analysis of cardiovesicle transcriptome from AS patients with respect to control patients in the VUMC cardiovascular cohort. **a.** Dot plots showing the expression of all “Bulk EV_unique” (EV_AS, 92) and “Cardiovesicle_unique” transcripts (POPDC2_AS and POPDC2_Ctr, 153) in the cardiomyocytes of each AS patient obtained using cardiac tissue single-nuclear RNA-sequencing demonstrating predominant cardiac specificity of “Cardiovesicle_unique” transcripts compared to “Bulk EV_unique” transcripts. **b.** Dot plots showing the expression of Bulk EV_unique and Cardiovesicle_unique transcripts in different heart cell types of each AS patient.

## Methods

## Acknowledgments

Research reported in this publication included work performed in the Integrated Mass Spectrometry Shared Resource supported by the National Cancer Institute of the National Institutes of Health under grant number P30CA033572. This work was supported by National Institutes of Health Common Fund grant UG3/UH3 TR002878 awarded to SD, TT and KJ as well as R35 HL150807 to SD. PG is supported by the American Heart Association Postdoctoral Fellowship (23POST1014230). BL is supported by investigator-initiated grant support from Edwards Lifesciences and consulting fees from Edwards Lifesciences and Astra Zeneca. ES is supported by National Heart, Lung and Blood Institute, National Institutes of Health (R01-HL151838). ERG receives funding from National Institutes of Health grants (NIH/NHGRI R01HG011138 and NIH/NIGMS R01GM140287). We thank the MGH Microscopy Core for TEM imaging

## Contribution

KVKJ, RS, and SD conceptualized, initiated, and supervised the study. PP, Ritin S, JFV, KGM performed the proteomics-based analysis of iPSC-CM EVs. MS and GL developed, tested, and optimized the Exocapture MSB technique. MS and PG performed cardiovesicle immunocapture and RNA isolations for Alpha MHC Cre-driven Exomap1 mice and patients (MGH – discovery, validation, and VUMC cohorts). JJ provided the MGH cohort, supervised, wrote and reviewed the manuscript. BM and PG performed library preparation for exRNA-sequencing for MGH and VUMC cohorts, respectively. PG performed qPCR validation for mouse and MGH cohorts. PP, KGM, RS, and JFV performed the LC-MS and subsequent preliminary bioinformatic analysis, and EH and BM performed the computational analysis and experimental methods to identify POPDC2 and CHRNE as cardiac EV membrane proteins. QS performed RNA-sequencing analysis for MGH and VUMC cohorts. RP, MEC helped in generating the Alpha MHC cre-driven Exomap1 mouse and sample preparation. NCA performed Imagestream and Vav cre experiments. BRL, SE, TA, KVA, and YRS were involved in the adjudication, acquisition, and processing of myocardial tissue and plasma samples, establishing the VUMC patient cohort tissue-plasma sample bank. ERG, RP, MJB, XB, and MLL were involved in the myocardial tissue snRNA sequencing of the VUMC cohort. CL and CA were involved in patient sample collection, storage and preprocessing along with patient data for the MGH cohort. ND performed the F-MRPS experiments. PS, MGC, TJT, IG performed experiments for the study. MS, PG wrote and edited the initial manuscript draft. DV, MK, DT, and AS carefully reviewed and edited the manuscript. MR, KT, and PTE supervised, guided, and reviewed the manuscript writing process and provided feedback. SD and RS supervised, conceptualized, and guided the experimental portion, and wrote, reviewed, and edited the manuscript.

MS, PG, and GL are equal contributors listed as first co-authors. EH, BM and QS are equal contributors listed as second co-authors. KVKJ, RS, and SD are senior co-authors. SD is the corresponding author.

